# Hydrogen Sulfide (H₂S)-Producing Oral Bacteria May Protect Against COVID-19

**DOI:** 10.1101/2024.08.07.24311606

**Authors:** Meghalbahen Vaishnani, Anupama Modi, Kshipra Chauhan, Bhavin Parekh

## Abstract

COVID-19 mortality rates have varied dramatically across the globe. Yet the reasons behind these disparities remain poorly understood. While recent research has linked gut microbes to these variations, the role of oral bacteria, a main port of entry for the coronavirus, remains unexplored.We investigated the relationship between oral microbiota and COVID-19 mortality rates across eightcountries. Raw sequencing data of 16S rRNA regions from oral microbiota in 244 healthy subjects from eight countries were obtained from public databases. We employed a generalized linear model (GLM) to predict COVID-19 mortality rates using oral microbiota composition. GLM revealed that high abundances of hydrogen sulfide (H₂S)-producing bacteria, particularly Treponema, predicted low COVID-19 mortality rates with a markedly low p-value. Unsupervised clustering using a combination of LIGER and t-SNE yielded four oral microbiome "orotypes." Orotypes enriched in H₂S-producing bacteria coincided with lower mortality rates, while orotypes harboring Haemophilus or Rothia were associated with increased vulnerability. To validate our findings, we analyzed influenza mortality data from the same countries, observing similar protective trends. Our findings suggest that oral bacteria-produced H₂S may serve as a critical initial defense against SARS-CoV-2 infection.H₂S from oral bacteria may prevent infection through antiviral activity, blocking ACE2 receptors, suppressing cytokines, and boosting antioxidants. This highlights the oral microbiome’s role in COVID-19 outcomes and suggests new preventive and therapeutic strategies.

**Graphical Abstract:** 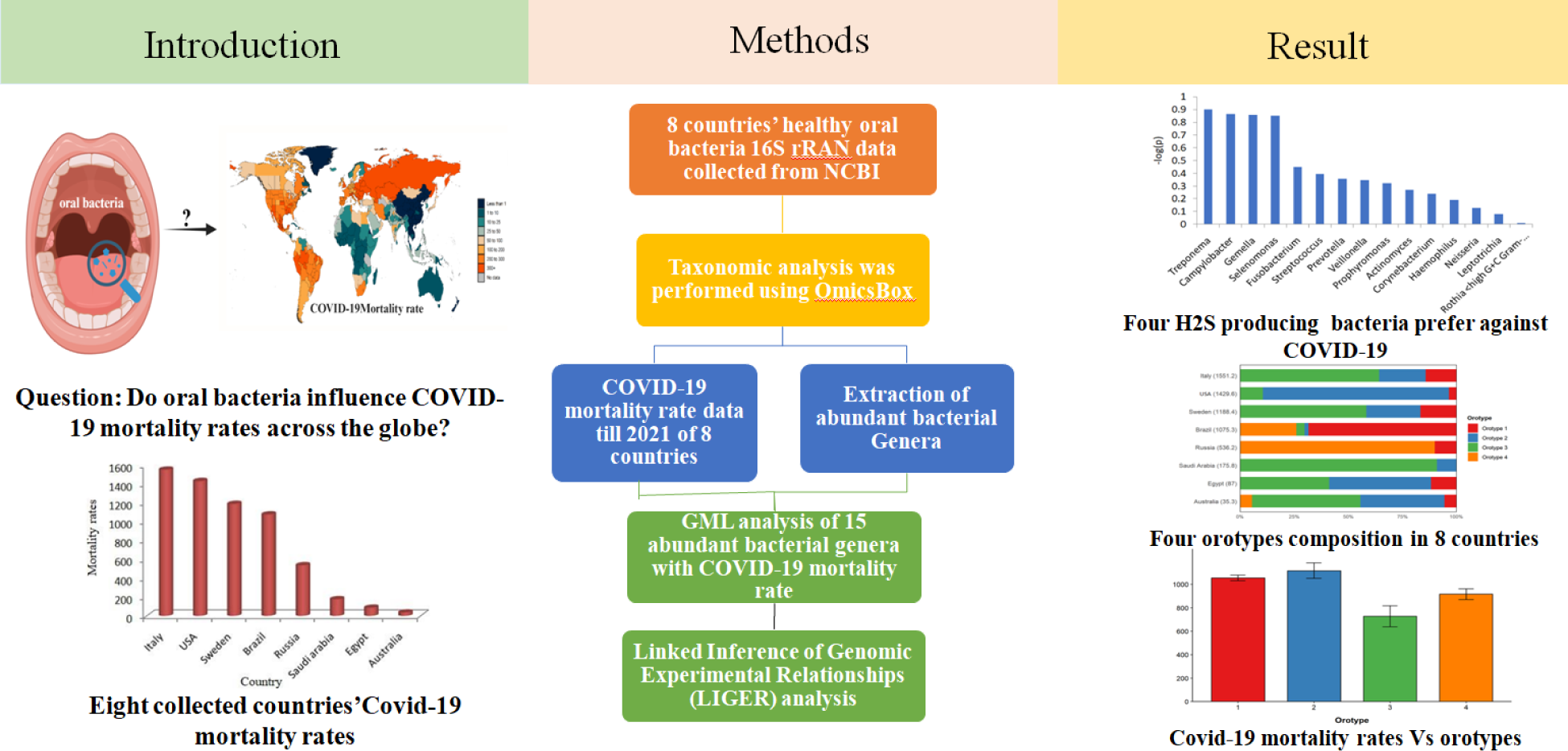

## Introduction

Why did COVID-19 hit some countries harder than others?[1]. This remains one of the pandemic’s most perplexing questions—a critical piece of the puzzle for future pandemic preparedness [1] [2]. Conventional risk factors like age and comorbidities like diabetes and obesity fail to fully explain this aspect [2] [3]. Emerging research implicates the human microbiome as a potential arbiter of COVID-19 outcomes [3]. Researchers have shown that low levels of certain gut bacteria, such as Collinsella, predict high COVID-19 mortality rates. Collinsella produces ursodeoxycholate, which may prevent infection and mitigate acute respiratory distress syndrome by suppressing cytokine storm syndrome[3].

What about the oral microbiota—the second largest microbial community in the body [4], SARS-CoV-2’s first contact point[5] and our frontline defense against respiratory viruses[6]?Exploring this overlooked aspect could further illuminate why some individuals succumb to the virus while others remain resilient[7]. Intrigued by this possibility, we hypothesize that dynamic interaction between the virus and specific oral microbiome may confer varying degrees of susceptibility or resilience to COVID-19 across populations.

To test this hypothesis, we analyzed the relationship between oral bacterial composition in 244 healthy subjects from eight countries and their respective COVID-19 mortality rates. Our findings reveal a negative correlation between four H_2_S-prodcing genera and COVID-19 mortality, suggesting a potential protective role for this oral commensal. To validate these findings, we further analyzed influenza mortality rates in the same countries, confirming similar protective trends and strengthening the generalizability of our results to respiratory viral infections.

## Materials and Methods

### Dataset

To investigate the potential association between oral microbiota composition and COVID-19 mortality rates, this study utilized publicly available healthy oral microbiome datasets from the National Center for Biotechnology Information (NCBI). The analysis encompassed data from eight countries: the United States, Australia, Brazil, Italy, Sweden, Saudi Arabia, Egypt, and Russia. These datasets were selected based on their accessibility and comprehensive coverage, corresponding to accession numbers PRJNA606501, PRJNA384402, PRJNA256234, PRJNA267483, PRJNA598825, PRJNA227796, PRJNA292800, and PRJDB5153. COVID-19 mortality data, quantified as cumulative deaths per million population as of February 9, 2021, were obtained from ‘https://ourworldindata.org/’. This timeframe was chosen to precede the widespread distribution of vaccines, thereby minimizing potential confounding effects of vaccination campaigns on mortality rates. To maintain the integrity and breadth of the analysis, no filtering criteria based on age or sex were applied to the datasets. This approach preserved a diverse and representative subject range, allowing for a more comprehensive examination of the potential relationship between oral microbiota and COVID-19 mortality across different populations.

### Taxonomic Analysis of oral microbiota

To classify the oral microbiota, OmicsBox (version 3.0.29) developed by BioBam Bioinformatics, utilizing the Kraken2 tool, was employed. Kraken2 performs taxonomic classification by analyzing k-mers in DNA short reads and querying a comprehensive database of species-specific k-mer information. This approach allowed for thorough and detailed taxonomic profiling, covering both metabarcoding (16S rRNA gene) and metagenomic sequencing reads, thus ensuring robust and accurate taxonomic identification.

### Generalized Linear Model (GLM) Analysis

To investigate the relationship between oral microbiota composition and COVID-19 mortality, Generalized Linear Models (GLMs) were employed. The 15 most abundant genera served as predictors, with COVID-19 mortality as the outcome variable(Fig 1). Model selection was performed using Akaike Information Criterion (AIC), comparing Gaussian, Gamma, and Inverse Gaussian distributions. The optimal model (Gamma distribution) was used to identify significant associations between specific genera and mortality rates.

**Fig 1.**
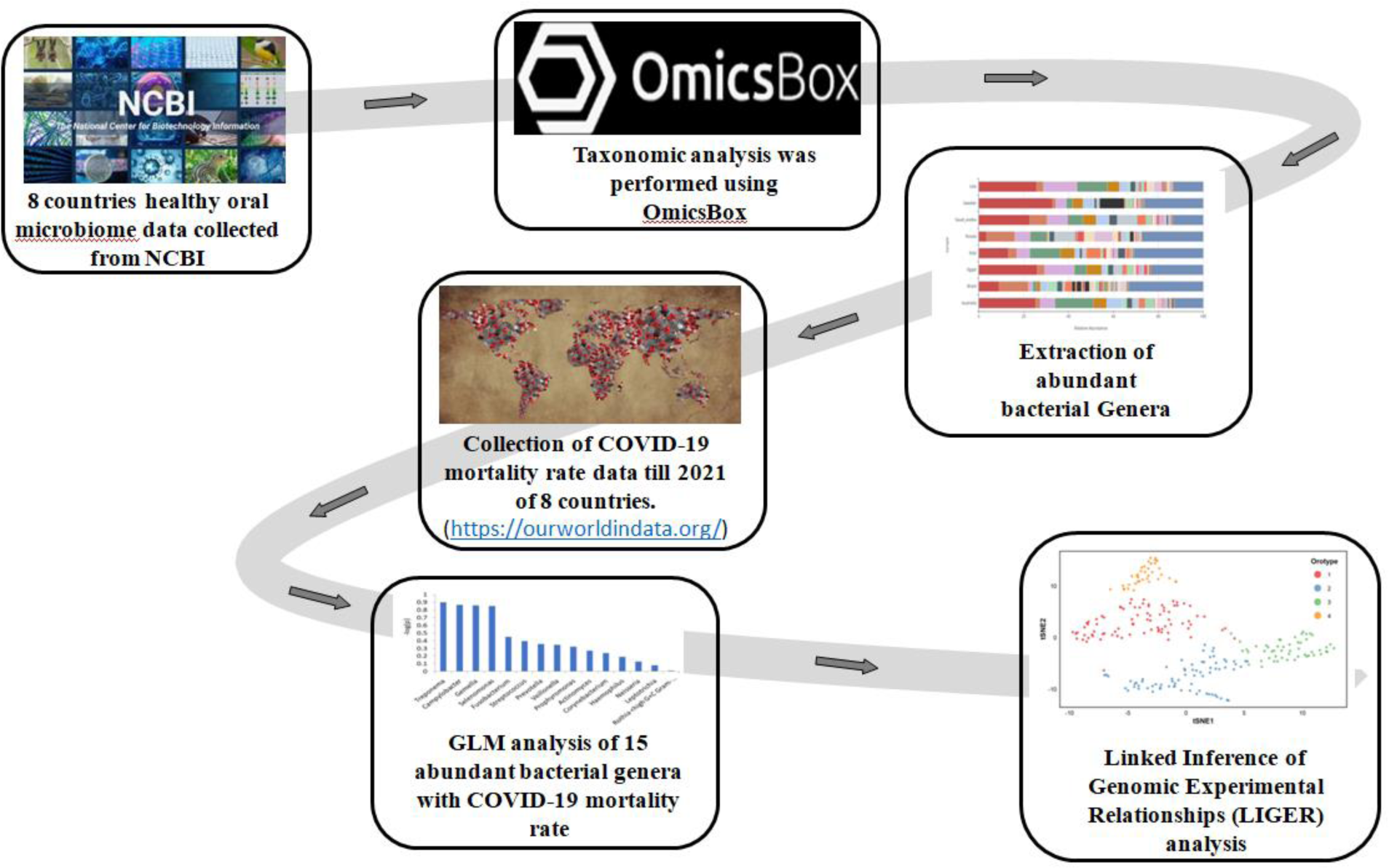
The methodology for this bioinformatics pipeline (a) Healthy oral microbiome data was obtained from the National Center for Biotechnology Information (NCBI). (b) This data was processed and analyzed using OmicsBox software. (c) The most abundant bacterial genera were identified. (d) COVID-19 mortality rate data from Our World in Data was incorporated (e) A statistical model (Generalized Linear Model) was used to analyze the relationship between the 15 most abundant genera and COVID-19 mortality. (f) A specialized technique called LIGER, typically used for analyzing single-cell RNA sequencing data, was applied to classify the oral microbiota of 244 healthy individuals into four distinct groups, termed "orotypes".

### LIGER-tSNE Analysis

A combination of Linked Inference of Genomic Experimental Relationships (LIGER) and t - distributed stochastic neighbor embedding (t-SNE) was employed in our analysis to explore and visualize the complex oral microbiome structures across different populations (Fig 1). This LIGER-tSNE combination was found to be a powerful tool for dimensionality reduction and clustering, allowing subtle patterns in the oral microbiome data that might have been overlooked by traditional analysis methods to be discerned. This innovative approach, originally developed for single-cell RNA sequencing data, proved highly effective in identifying distinct oral microbiome orotypes and their relationship to COVID-19 mortality rates.

## Results

### Generalized Linear Model (GLM) Analysis

16S rRNA sequencing data from 244 healthy subjects across eight countries (Table 1) were analyzed to examine the effects of oral microbiota on COVID-19 mortality rates. Relative abundance analysis at the genus level (Fig 2) revealed *Streptococcus, Prevotella, Veillonella, Fusobacterium*, and *Haemophilus* as the most prevalent genera.

**Fig 2:**
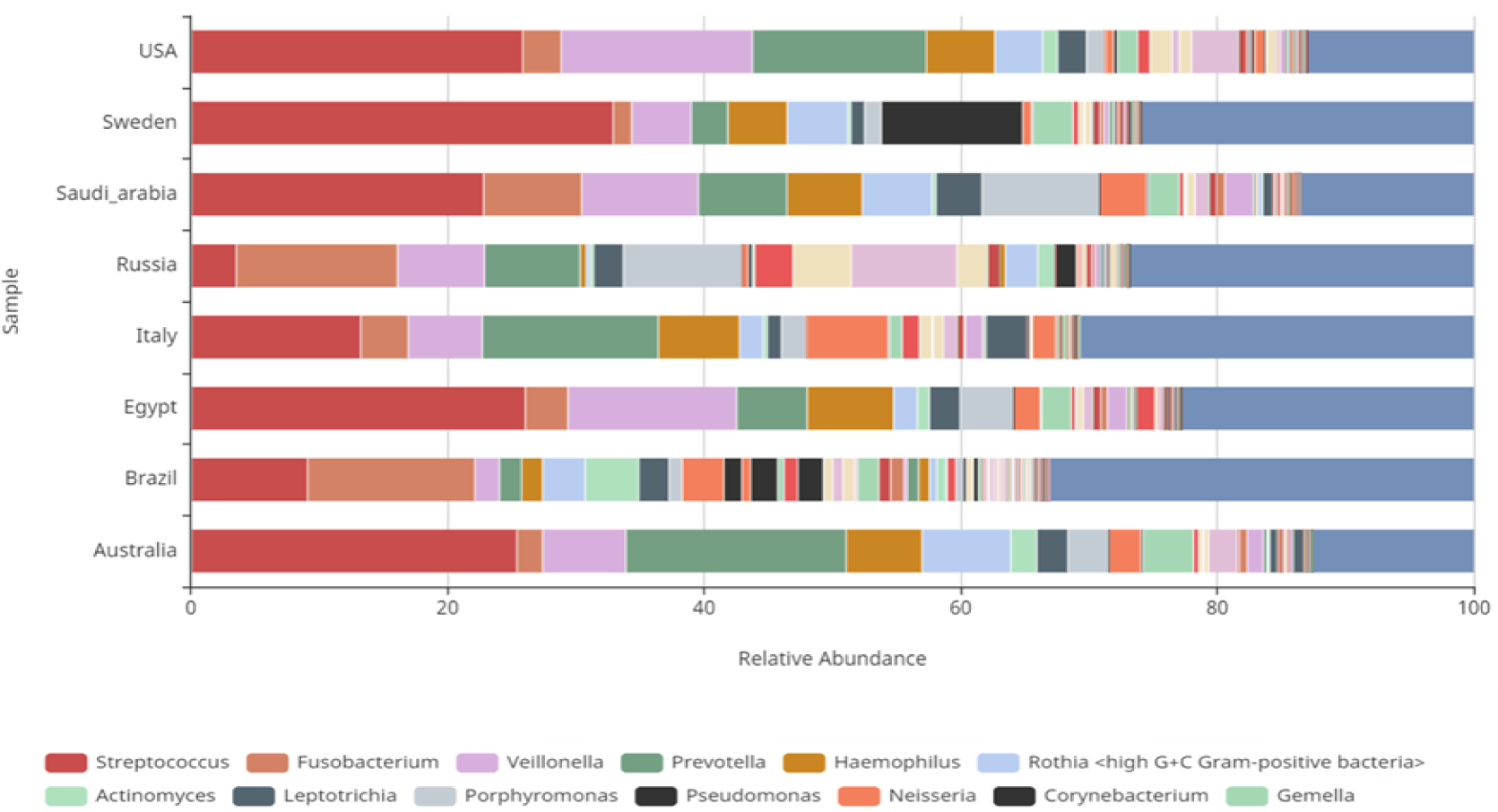
Genera level abundance of health oral metagenomic

**Table 1:**
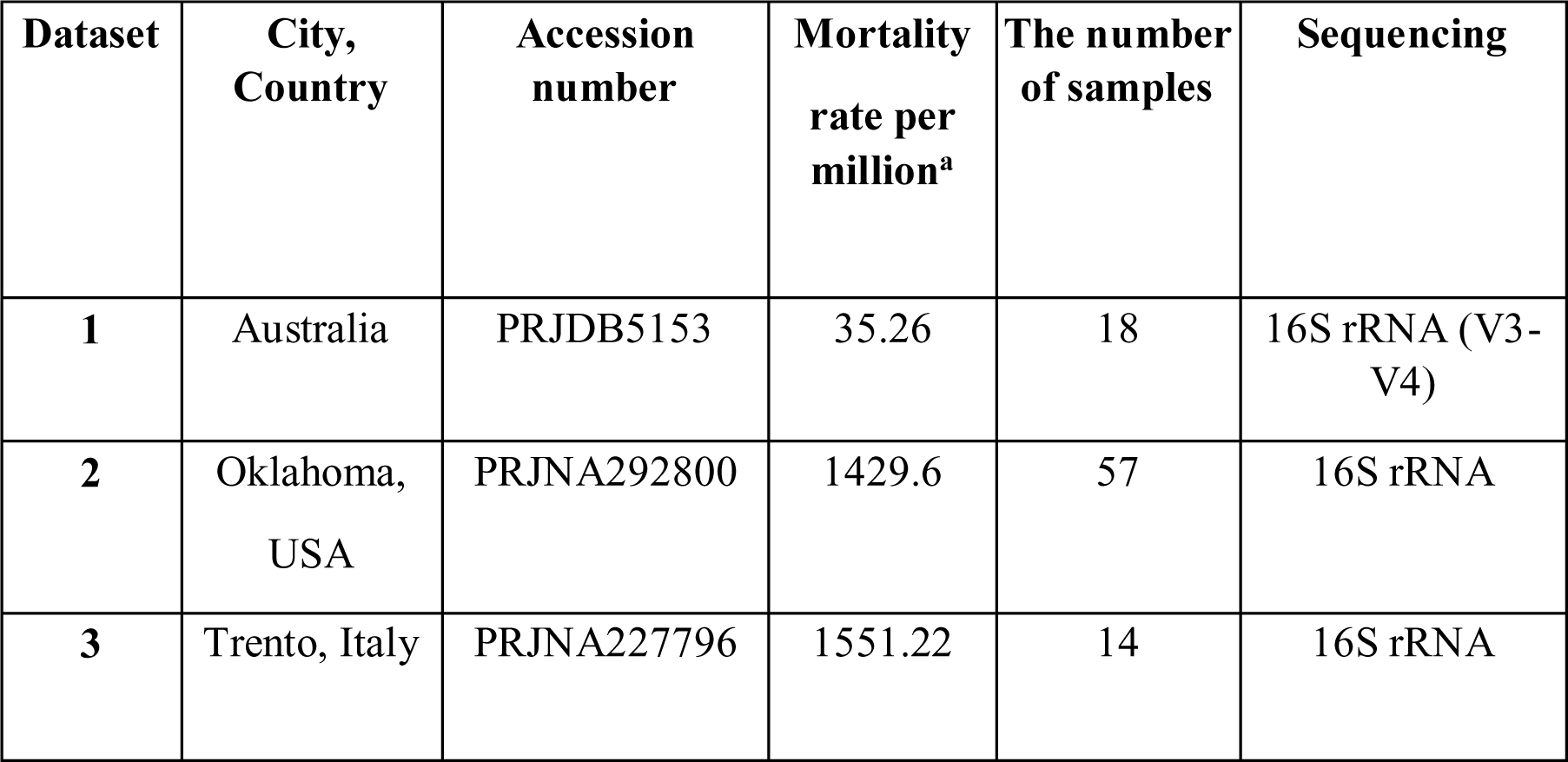
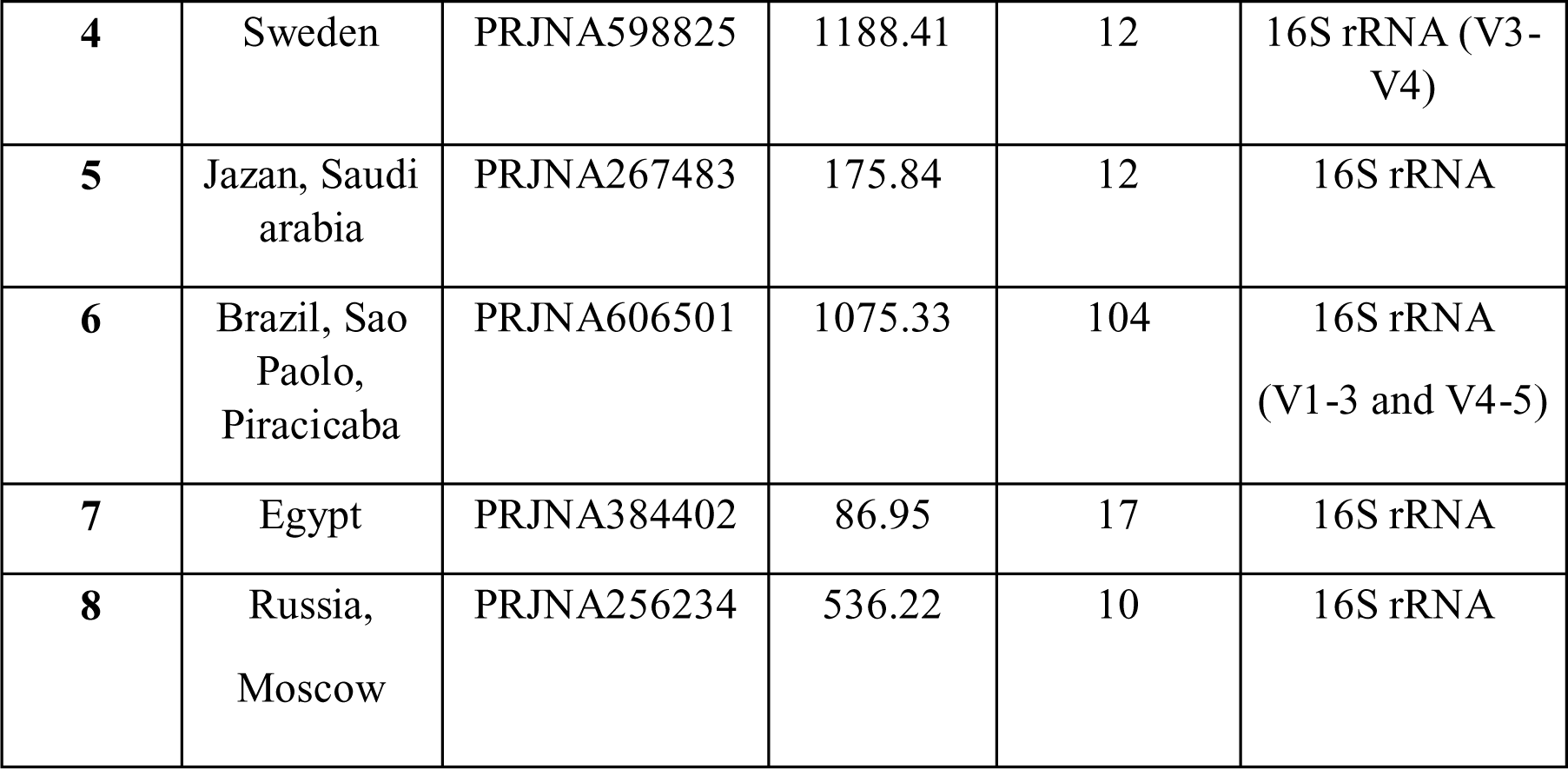
Eight 16S rRNA-seq datasets from eight countries and the mortality rates of COVID-19.

| Dataset | City, Country | Accession number | Mortality rate per million<sup>a</sup> | The number of samples | Sequencing |
| --- | --- | --- | --- | --- | --- |
| 1 | Australia | PRJDB5153 | 35.26 | 18 | 16S rRNA (V3-V4) |
| 2 | Oklahoma, USA | PRJNA292800 | 1429.6 | 57 | 16S rRNA |
| 3 | Trento, Italy | PRJNA227796 | 1551.22 | 14 | 16S rRNA |
| **4** | Sweden | PRJNA598825 | 1188.41 | 12 | 16S rRNA (V3-V4) |
| **5** | Jazan, Saudi arabia | PRJNA267483 | 175.84 | 12 | 16S rRNA |
| **6** | Brazil, Sao Paolo, Piracicaba | PRJNA606501 | 1075.33 | 104 | 16S rRNA (V1-3 and V4-5) |
| **7** | Egypt | PRJNA384402 | 86.95 | 17 | 16S rRNA |
| **8** | Russia, Moscow | PRJNA256234 | 536.22 | 10 | 16S rRNA |

| **Dataset** | **City, Country** | **Accession number** | **Mortality rate per million<sup>a</sup>** | **The number of samples** | **Sequencing** |
| --- | --- | --- | --- | --- | --- |
| **1** | Australia | PRJDB5153 | 35.26 | 18 | 16S rRNA (V3-V4) |
| **2** | Oklahoma, USA | PRJNA292800 | 1429.6 | 57 | 16S rRNA |
| **3** | Trento, Italy | PRJNA227796 | 1551.22 | 14 | 16S rRNA |
| **4** | Sweden | PRJNA598825 | 1188.41 | 12 | 16S rRNA (V3-V4) |
| **5** | Jazan, Saudi arabia | PRJNA267483 | 175.84 | 12 | 16S rRNA |
| **6** | Brazil, Sao Paolo, Piracicaba | PRJNA606501 | 1075.33 | 104 | 16S rRNA<br>(V1-3 and V4-5) |
| **7** | Egypt | PRJNA384402 | 86.95 | 17 | 16S rRNA |
| **8** | Russia, Moscow | PRJNA256234 | 536.22 | 10 | 16S rRNA |

Using GLM, COVID-19 mortality rates were predicted with the 15 most abundant genera, with the Gamma distribution yielding the lowest AIC. GLM results (Fig 3) highlighted a marked negative predictive value of *Treponema* abundance for COVID-19 mortality rates (p < 0.001). *Campylobacter, Gemella, and Selenomonas* also showed negative correlations, albeit less significant.

**Fig 3:**
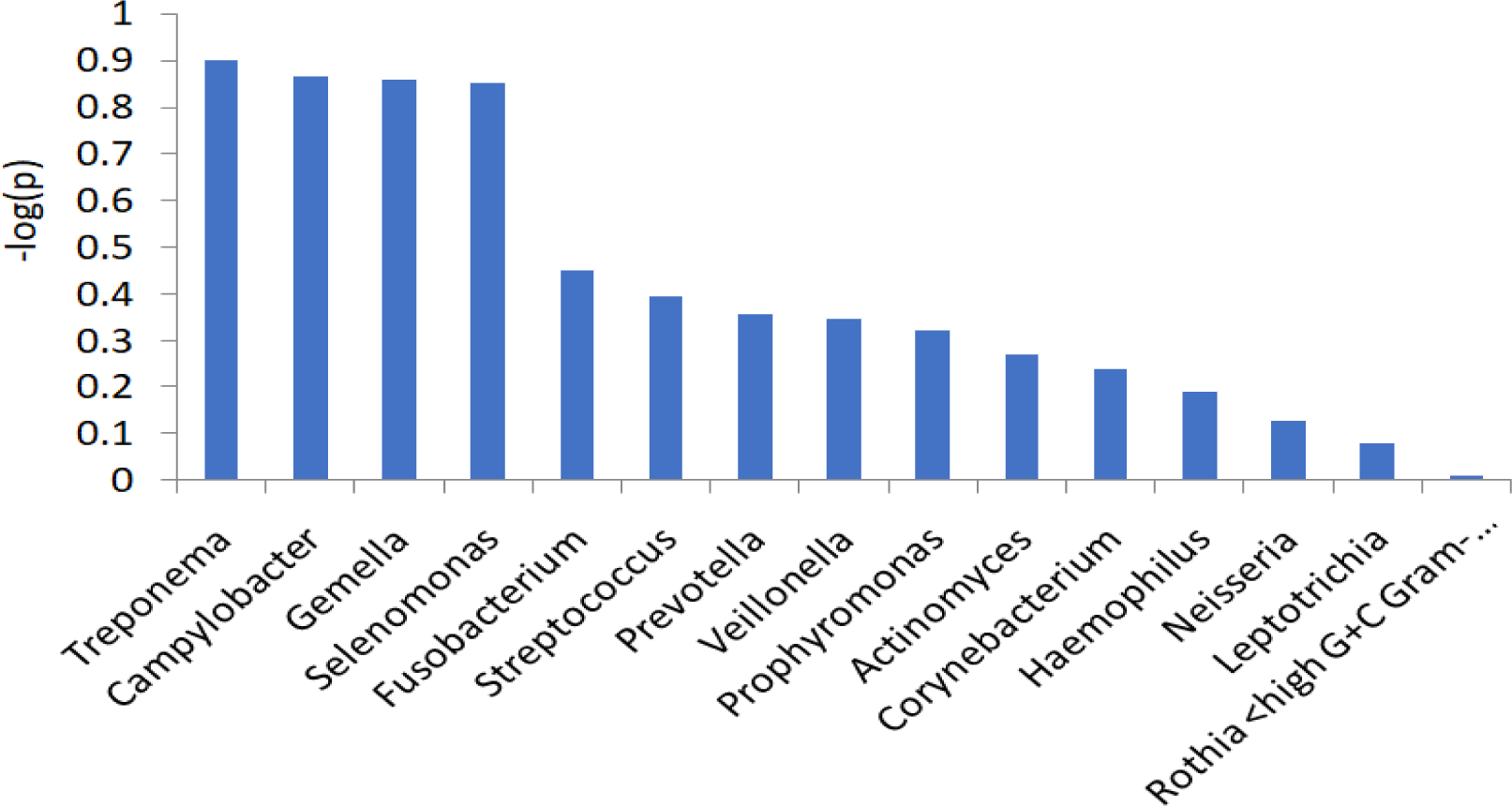
Plot of p-values of 15 genera in a generalized linear model (GLM) to predict the COVID-19 mortality rates.

### Linked Inference of Genomic Experimental Relationships (LIGER) Analysis

Non-negative matrix factorization via LIGER identified four distinct oral microbiome clusters, termed "orotypes" (Fig 4A). For every orotype, the mean relative abundances of the top 15 genera are calculated in (S3 Table). The most abundant 5 genera in each orotype are given in Supplementary Table 1(S2 Table).. orotype distribution varied across countries in relation to COVID-19 mortality rates (Fig 4B). High mortality countries (e.g., Italy, USA) predominantly exhibited orotypes 2 and 3, while low mortality countries (e.g., Australia, Russia) showed a higher prevalence of orotype 4 (Fig 4B).Countries with intermediate mortality rates displayed a mixed distribution.Analysis of COVID-19 mortality rates across orotypes (Fig 4C) revealed a decreasing trend from orotypes 1 to 4, although not statistically significant (p = 0.203, Jonckheere-Terpstra trend test, Fig 4D). Notably, *Treponema* relative abundance significantly increased from orotypes 1 to 4 (Fig 4E; p = 0.033, Jonckheere-Terpstra trend test, Fig 4F).

**Fig 4.**
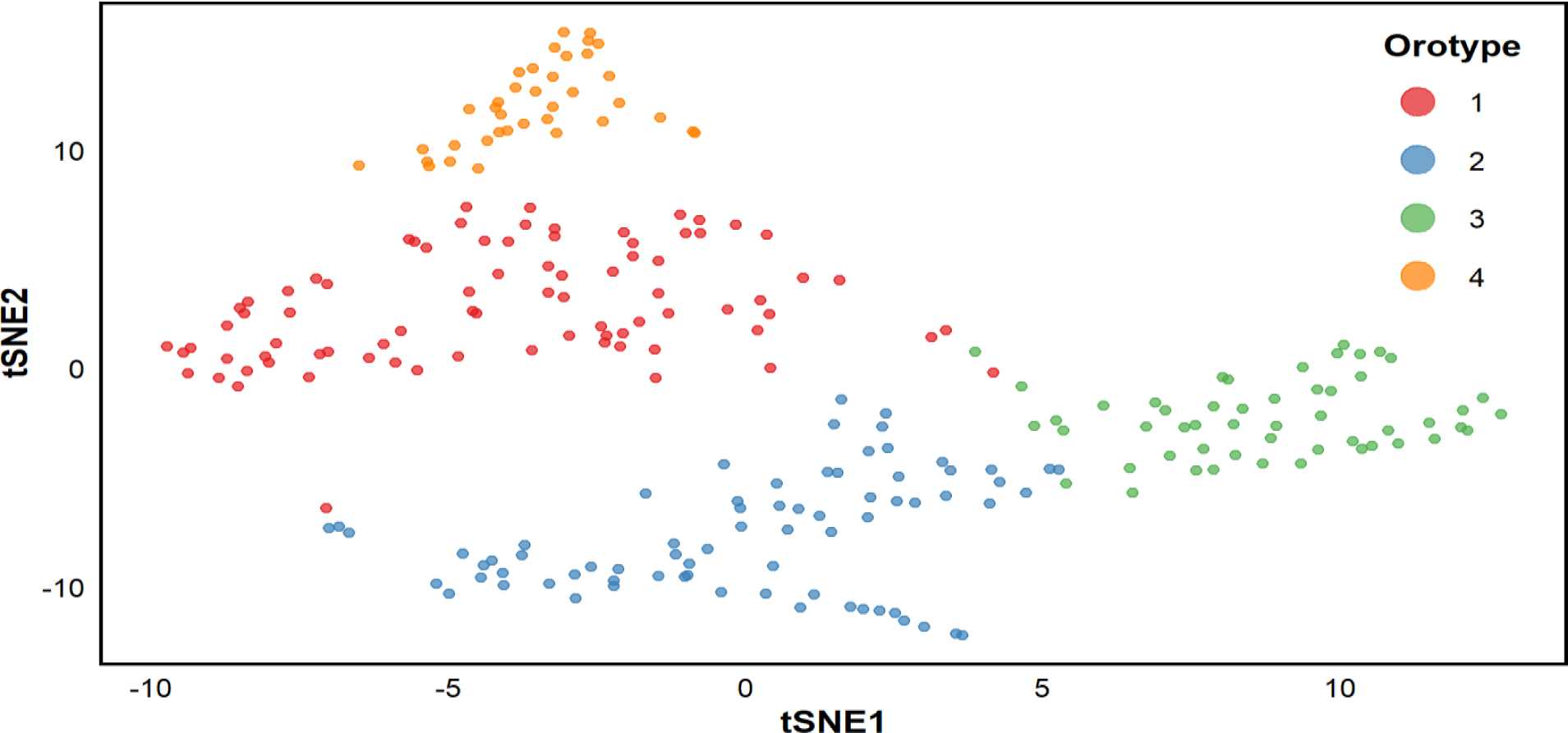
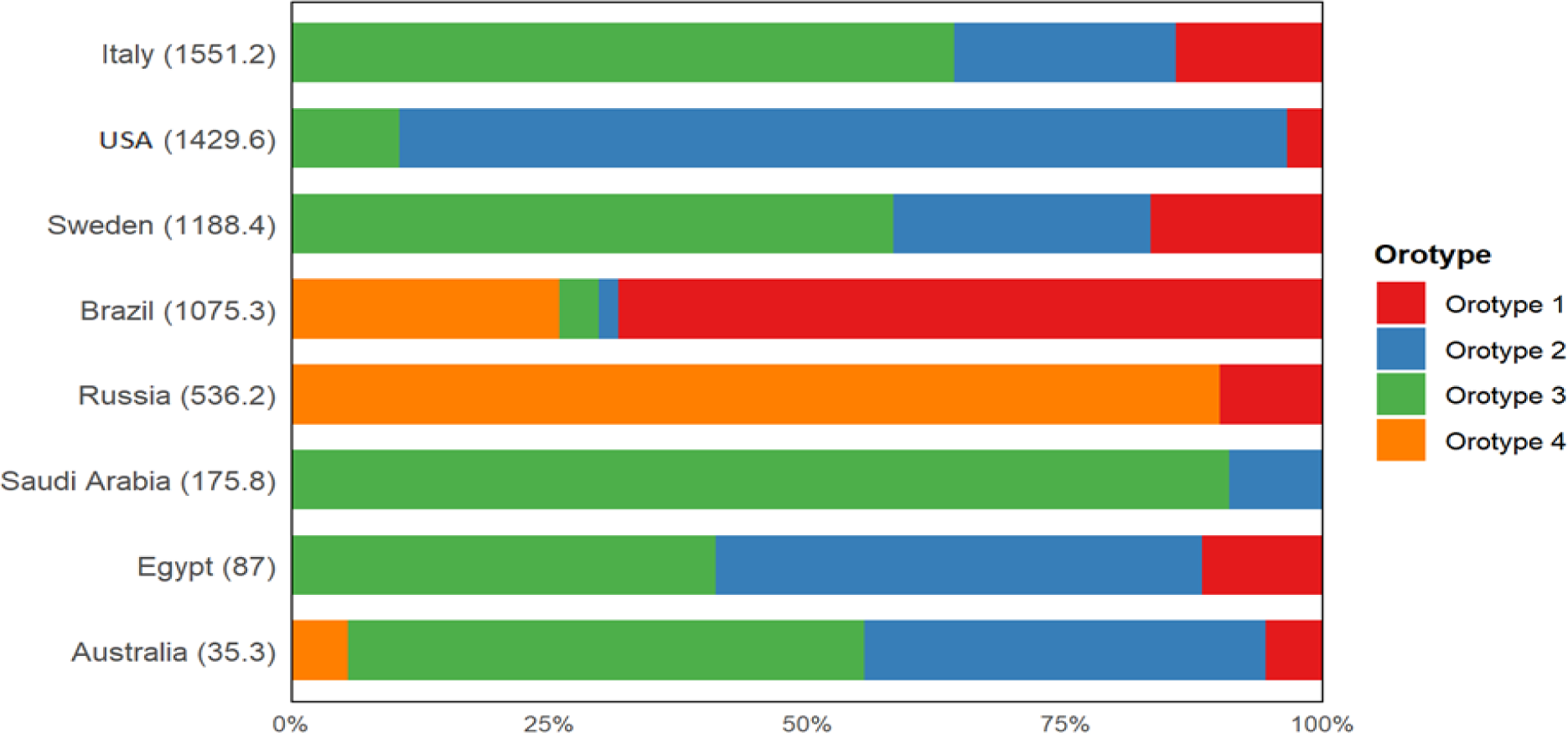
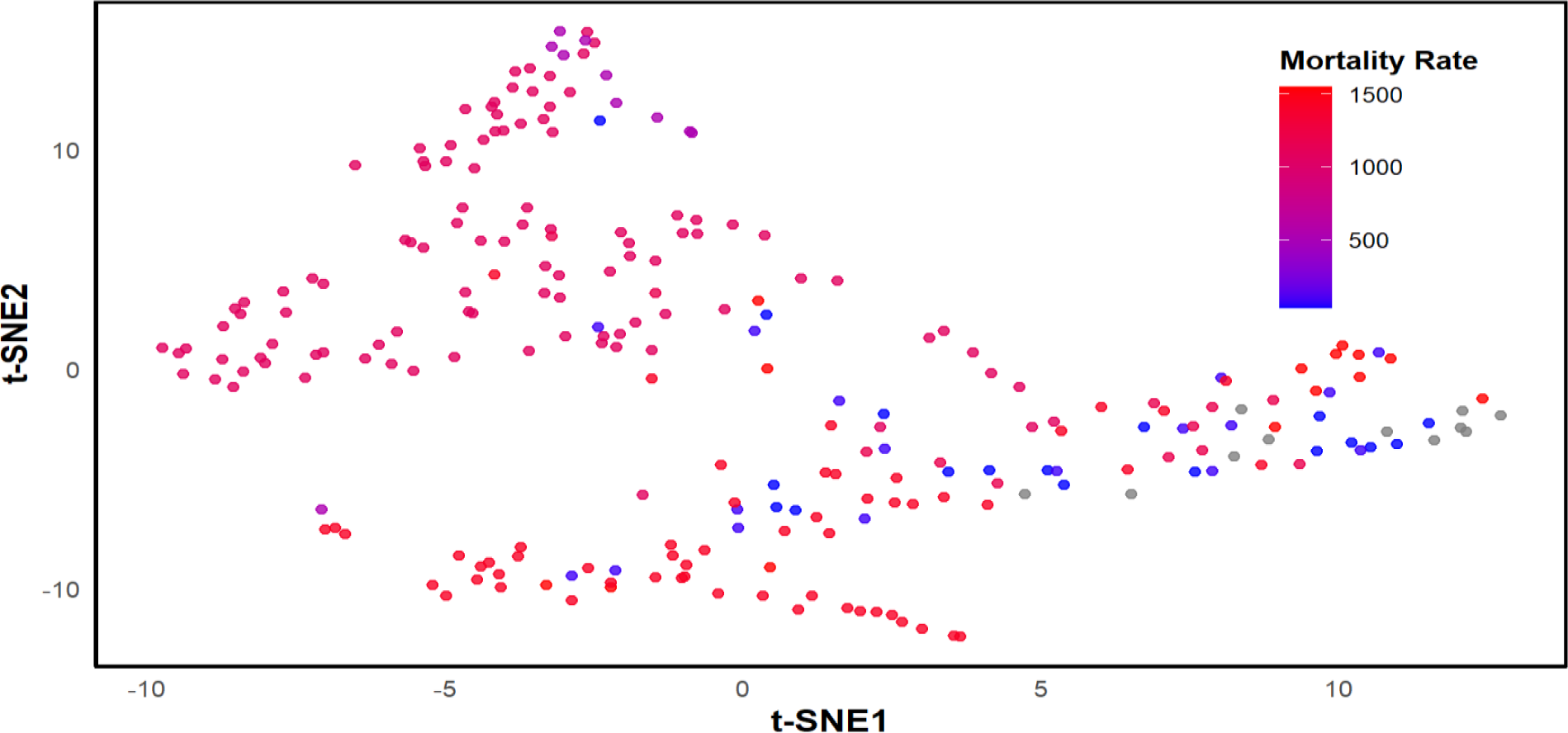
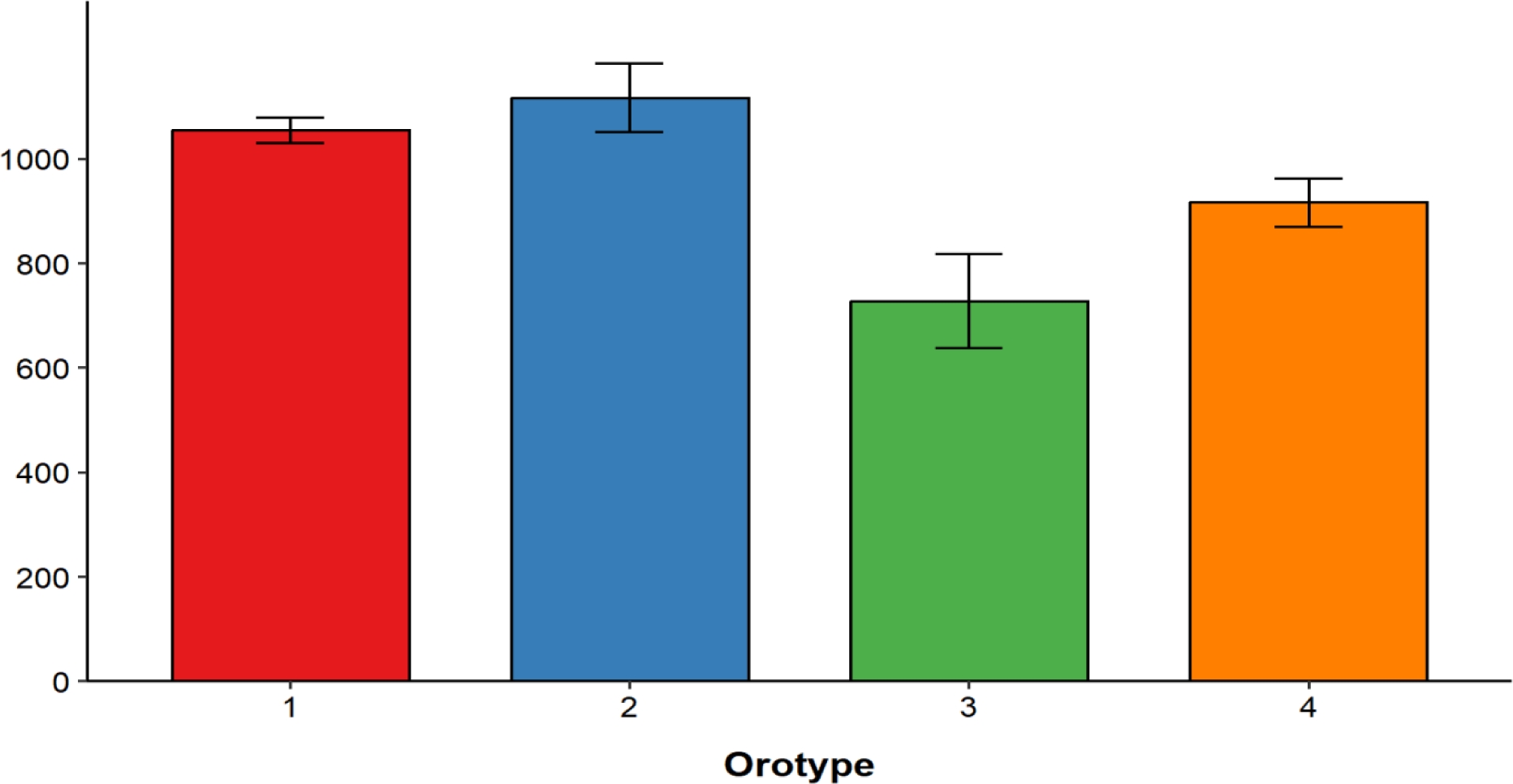
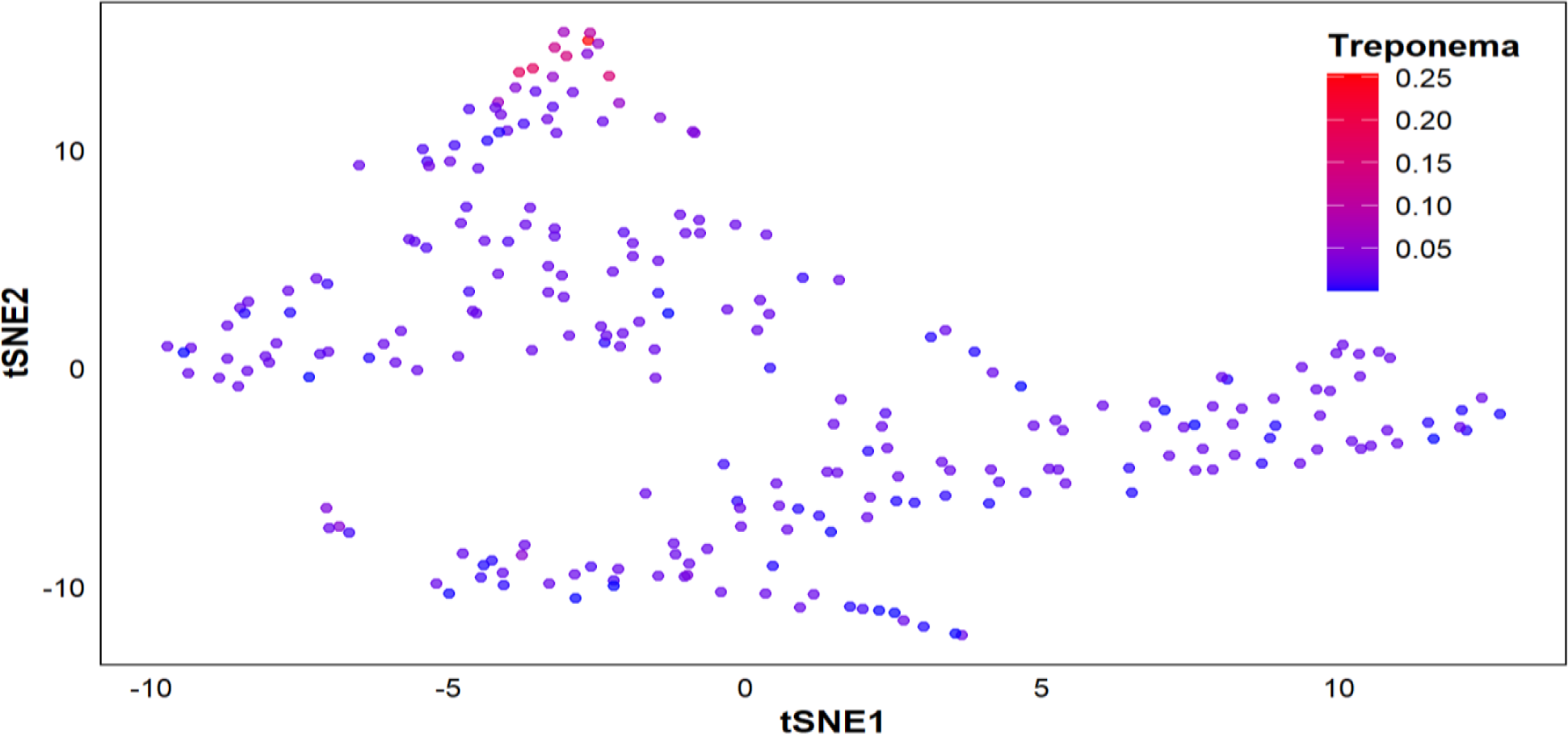
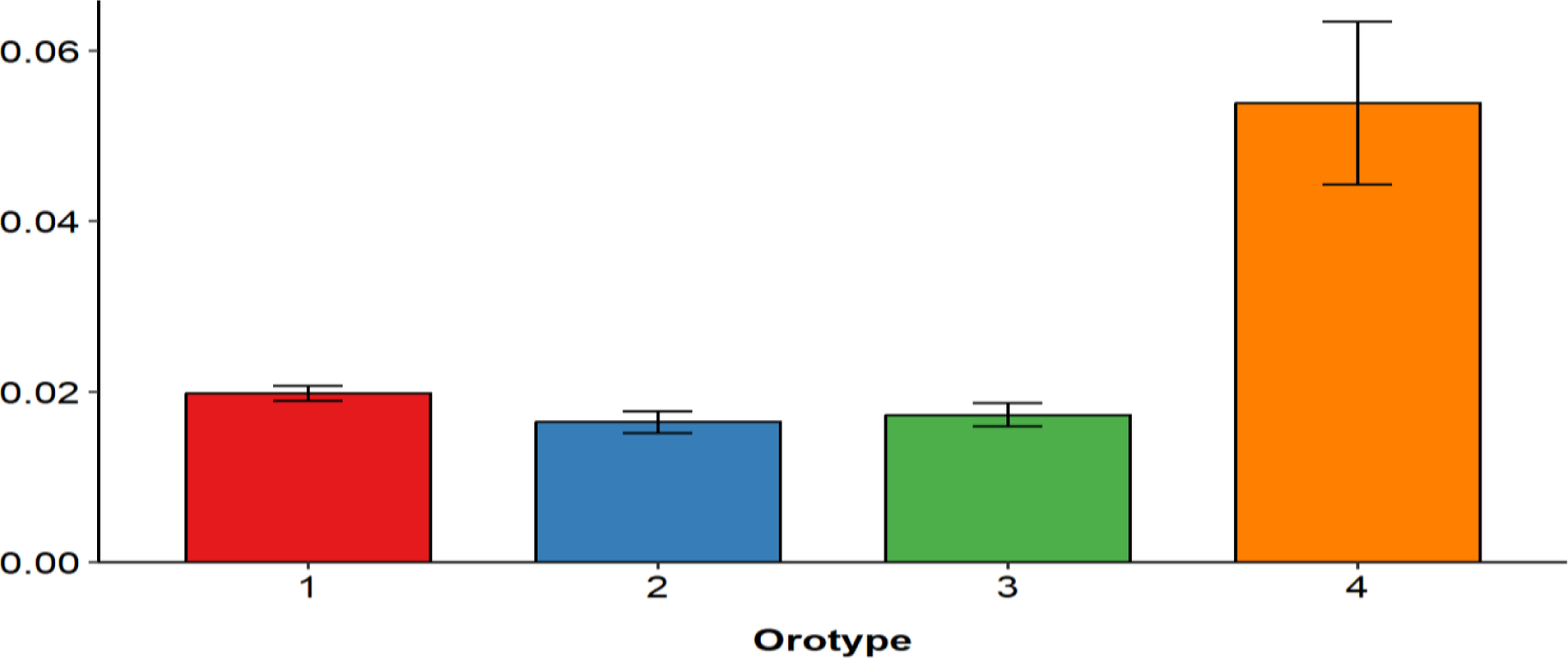
(a) Unsupervised clustering of oral microbiota in 244 healthy subjects in ten countries by LIGER generated 4 orotypes. Each subject is plotted with t-SNE and is color-coded by its orotype. (b) Fractions of orotypes 1 to 4 in eight countries. eight countries are sorted in descending order of the COVID-19 mortality rates per million, which are indicated in parentheses. (c) The t-SNE plot is color-coded by the COVID-19 mortality rates in eight countries. (d) Mean and standard error of the COVID-19 mortality rates in orotypes 1 to 4. *p = 0.203* by Jonckheere-Terpstra trend test. (e) The t-SNE plot is color-coded by the relative abundance of genus *Treponema*. (f) Mean and standard error of the relative abundance of genus *Treponema* in orotypes 1 to 4. *P* = 0.033by Jonckheere-Terpstra trend test.

### Other Genera of Interest and Combined Analysis

Analysis of Campylobacter, Gemella, and Selenomonas individually revealed no significant trends across orotypes (Figs S1a-f). However, combined analysis of Treponema, Gemella, Campylobacter, and Selenomonas demonstrated a highly significant increasing trend from orotypes 1 to 4 (p = 7.198e-05, Jonckheere-Terpstra trend test, Figs S1g-h), suggesting a potential synergistic effect of these bacteria.

### Validation with Influenza Mortality Data

To validate these findings, influenza mortality rates in these eight countries (taken from ‘https://ourworldindata.org/’ (2019)) were analyzed, given the similarities between COVID-19 and influenza as respiratory viral illnesses. The influenza mortality chart (Fig S2) demonstrates that orotype 4 is linked to lower mortality rates in influenza, mirroring the trends observed with COVID-19. This further substantiates the potential protective role of orotype 4-associated microbial composition in respiratory viral infections.

## Discussion

This study uncovers a fascinating link between specific oral bacteria—Treponema, Gemella, Campylobacter, and Selenomonas—and lower COVID-19 mortality rates (Fig 3). The unifying characteristic of these bacteria is their production of hydrogen sulfide (H₂S), a molecule now implicated as a pivotal protective agent against respiratory viral infections. Our data reveal a striking pattern in oral microbial communities. Those dominated by Haemophilus (orotype 1) or Rothia (orotype2) appear to be associated with increased vulnerability to SARS-CoV-2 (S2 Table). In contrast, communities enriched with H₂S-producing bacteria (orotypes 3 and 4) exhibit a protective effect.

This pattern echoes recent findings by Ren et al., who observed a similar association between Gemella and Rothia abundance in the upper respiratory tract of healthy individuals and COVID-19 patients, respectively. Wu et al. (2021) also reported significant microbiome alterations in COVID-19 patients, including elevated Rothia mucilaginosa levels in both oral and gut samples. Interestingly, this potential protective role of H₂S aligns with previous research on gut microbiota, where Collinsella, another H₂S-producing bacterium, was identified as a protective factor against COVID-19 [3] [8].

The H₂S-producing capacity of the identified bacteria is notable. Treponema species, particularly Treponema denticola, are notable for their high H₂S production capacity due to multiple cysteine-degrading enzymes, potentially explaining their strong negative correlation with COVID-19 mortality[9] [10].Gemella species such as Gemella morbillorum, while less studied in this context, has been identified as a H₂S producer, albeit at lower rates compared to some other oral bacteria[9]. Campylobacter species, particularly C. rectus, and Selenomonas thrive in anaerobic conditions and are known H₂S producers[9].

But how exactly does H₂S exert its protective effects?Its antiviral properties, previously demonstrated against other RNA viruses including influenza and respiratory syncytial virus (RSV) infections [11] [12], likely extend to SARS-CoV-2. These include direct antiviral activity[12], modulation of viral entry via ACE2 and TMPRSS2 receptors[13], (Fig 5) anti-inflammatory effects[12], prevention of cytokine storm[14],and enhancement of antioxidant defenses through the Nrf2 pathway [15] (Fig-5). Our findings are consistent with recent clinical observations showing higher serum H₂S levels in COVID-19 pneumonia survivors [16], reinforcing its potential as both a prognostic marker and therapeutic target. Furthermore, the validation of our results using influenza mortality data strengthens the broader implications of our findings, suggesting that the protective effect of H_2_S producing bacteria extends beyond COVID-19 to other respiratory viral infections.

**Fig 5:**
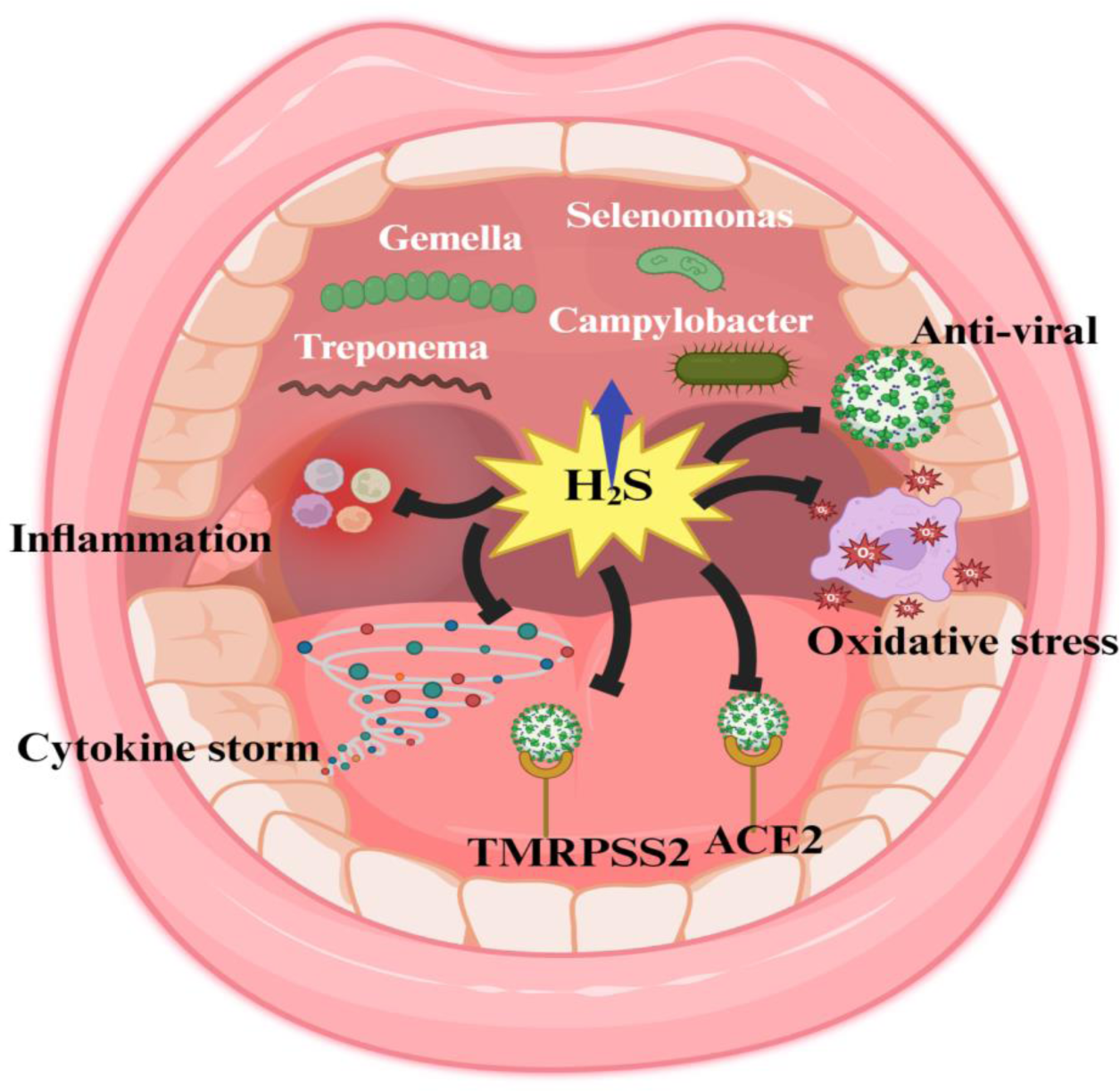
Proposed mechanisms of oral bacteria-produced hydrogen sulfide (H₂S) in protecting against SARS-CoV-2 infection. H₂S, produced by oral bacteria including Treponema, Gemella, Campylobacter, and Selenomonas, exhibits multifaceted protective effects against SARS-CoV-2. These include: (a) Direct antiviral activity, (b) Modulation of viral entry by altering ACE2 and TMPRSS2 receptors, (c) Anti-inflammatory effects and prevention of cytokine storm, and (d) Enhancement of antioxidant defenses through the Nrf2 pathway. These mechanisms, previously observed in other RNA viral infections, likely contribute to the potential protective role of H₂S-producing oral bacteria against severe COVID-19 outcomes.

Despite our promising findings, our study has several limitations. The observational nature of this study precludes establishing causality. Reliance on existing datasets may introduce biases, and the precise mechanisms of H₂S protection warrant further investigation. Additionally, individual variations in oral hygiene and diet could not be controlled. These limitations highlight the need for longitudinal studies, controlled experiments, and clinical trials.

In conclusion, our study identifies a novel association between H₂S-producing oral bacteria and reduced COVID-19 mortality, underscoring the oral microbiome’s potential role in modulating respiratory viral infection outcomes. This work paves the way for innovative approaches in the fight against respiratory viral diseases, including the exploration of H₂S-based therapeutics and probiotics.

## Supporting information

Fig S1a: The t-SNE plot is color-coded by the relative abundance of genus Campylobacter.

Fig S1b: Mean and standard error of the relative abundance of genus Campylobacter in orotypes 1 to 4. P = 0.236 by Jonckheere-Terpstra trend test.

Fig S1c: The t-SNE plot is color-coded by the relative abundance of genus Gemella.

Fig S1d: Mean and standard error of the relative abundance of genus Gemella in orotypes 1 to 4. P = 0.652 by Jonckheere-Terpstra trend test.

Fig S1e: The t-SNE plot is color-coded by the relative abundance of genus Selenomonas.

Fig S1f: Mean and standard error of the relative abundance of genus Selenomona in orotypes 1 to 4. P = 0.859 by Jonckheere-Terpstra trend test.

Fig S1g: The t-SNE plot is color-coded by the relative combine abundance of genus Treponema, Gemella, Campylobacter, Selenomonas.

Fig S1h: Mean and standard error of the relative combine abundance of genus Treponema, Gemella, Campylobacter, Selenomonas in orotypes 1 to 4. p = 7.

Fig S2: Mean and standard error of the Influenza mortality rates in orotypes 1 to 4. p = 4e - 04 by Jonckheere-Terpstra trend test.

Supplementary Table 1: Generalized linear model (GLM) to predict the COVID-19 mortality rates with 15 oral bacteria

Supplementary Table 2: Most abundance genera in each orotype

Supplementary Table 3: The mean relative abundances of 15 most prevalent genera for each orotype

## Data Availability

All data produced are available online at

https://www.ncbi.nlm.nih.gov/

https://ourworldindata.org/

## Abbreviation

COVID-19: Coronavirus disease 2019
SARS-CoV-2: Severe Acute Respiratory Syndrome Coronavirus 2
GLM: Generalized linear model
LIGER: Linked Inference of Genomic Experimental Relationships
H_2_S: Hydrogen sulfide
t-SNE: t-distributed Stochastic Neighbor Embedding
NCBI: National Center for Biotechnology Information
RNA: Ribonucleic Acid
DNA: Deoxyribonucleic acid
RSV: Respiratory syncytial virus
Nrf2: nuclear factor erythroid 2–related factor 2

## Acknowledgement

None

## Disclosure of interest

**No** potential conflict was reported by the anthor(s)

## Funding

None

## Authour Contribution

BP conceptualized the study, performed data analysis, interpreted the data, and wrote the manuscript. MV conducted data analysis, participated in the creation of graphics, tables, and charts, and managed the submission process. AM reviewed, edited, and refined the manuscript, and coordinated the submission process. KC assisted with initial data collection efforts and offered input on the taxonomic analysis approach.

## Supplementary Information

**Fig S1:**
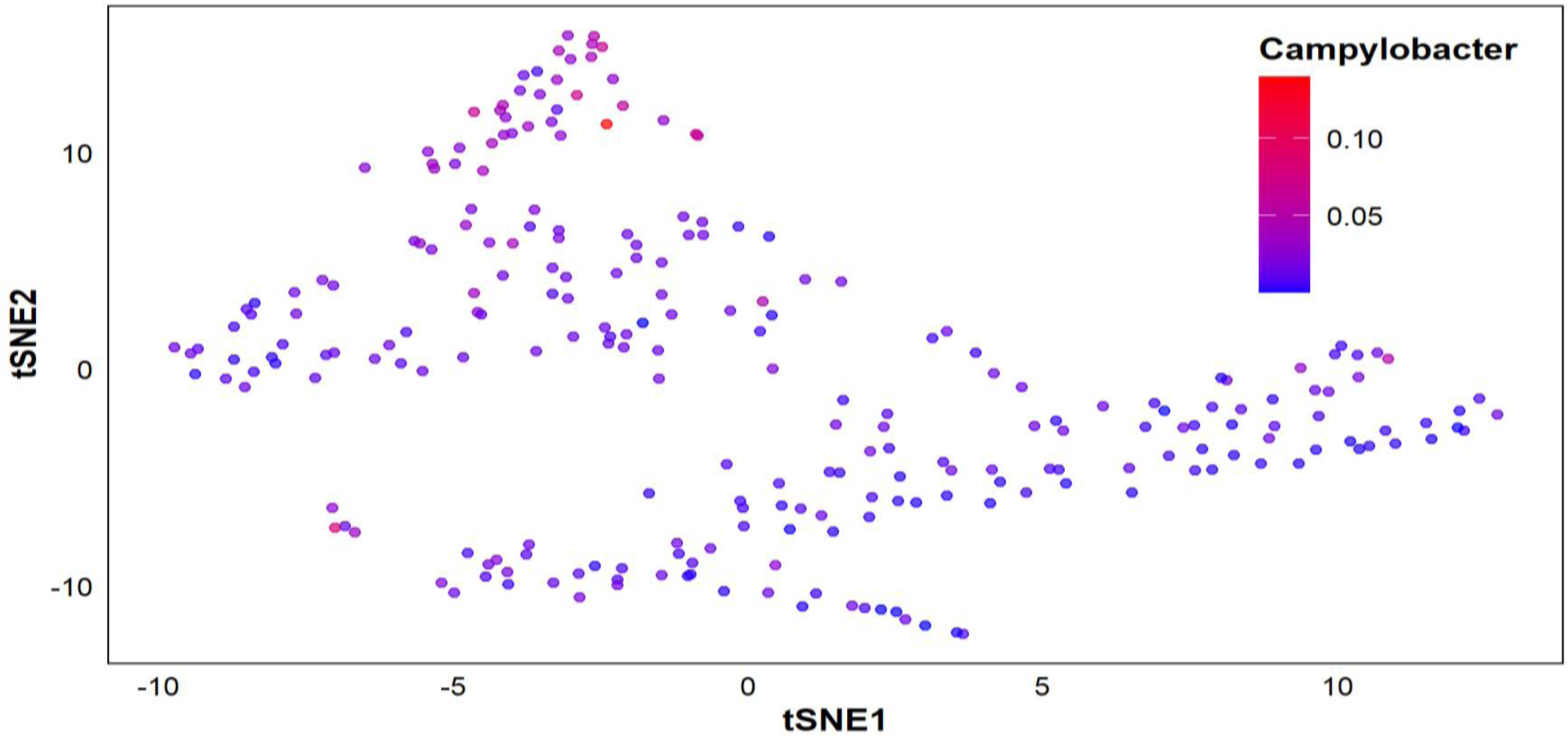
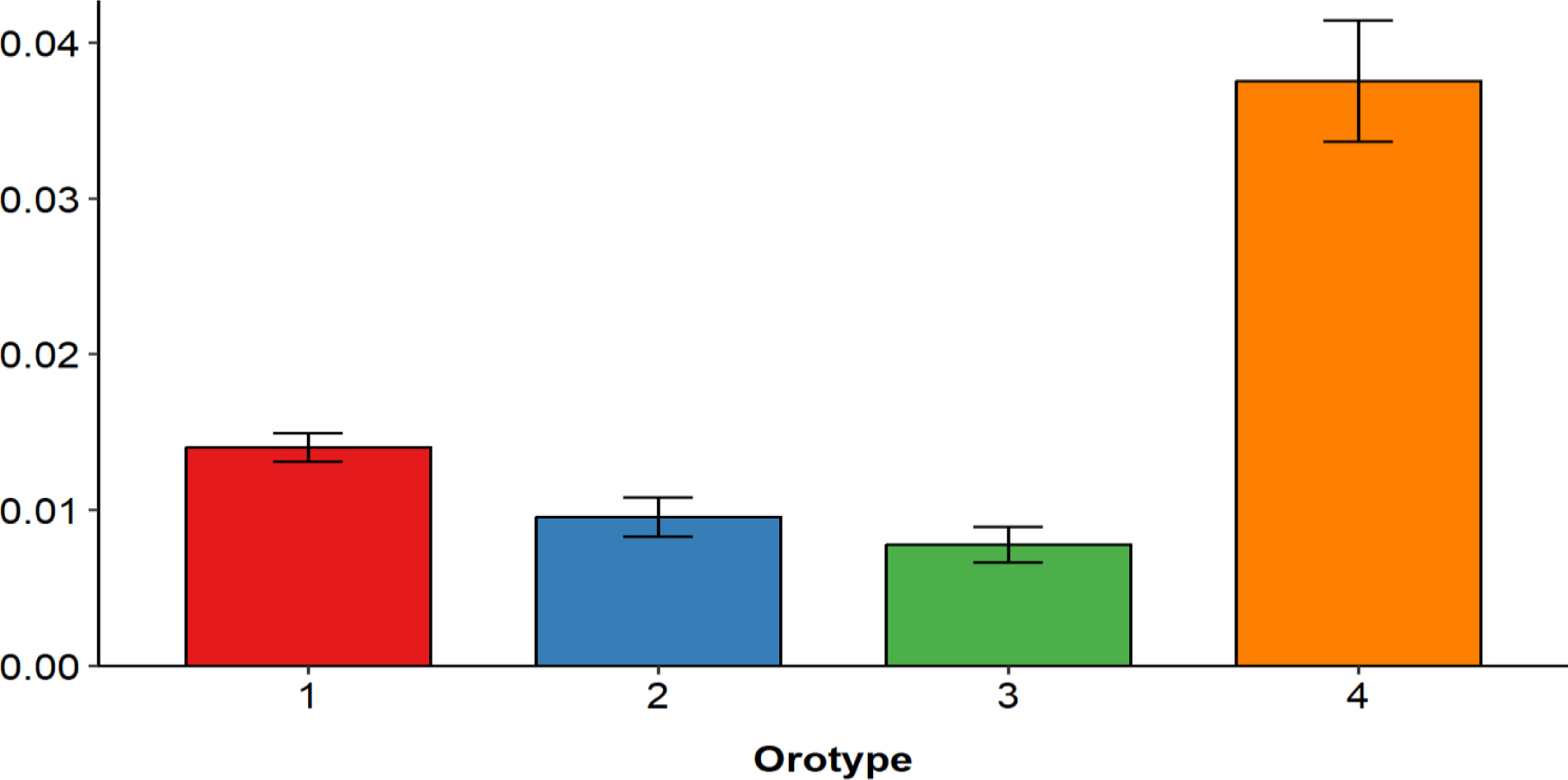
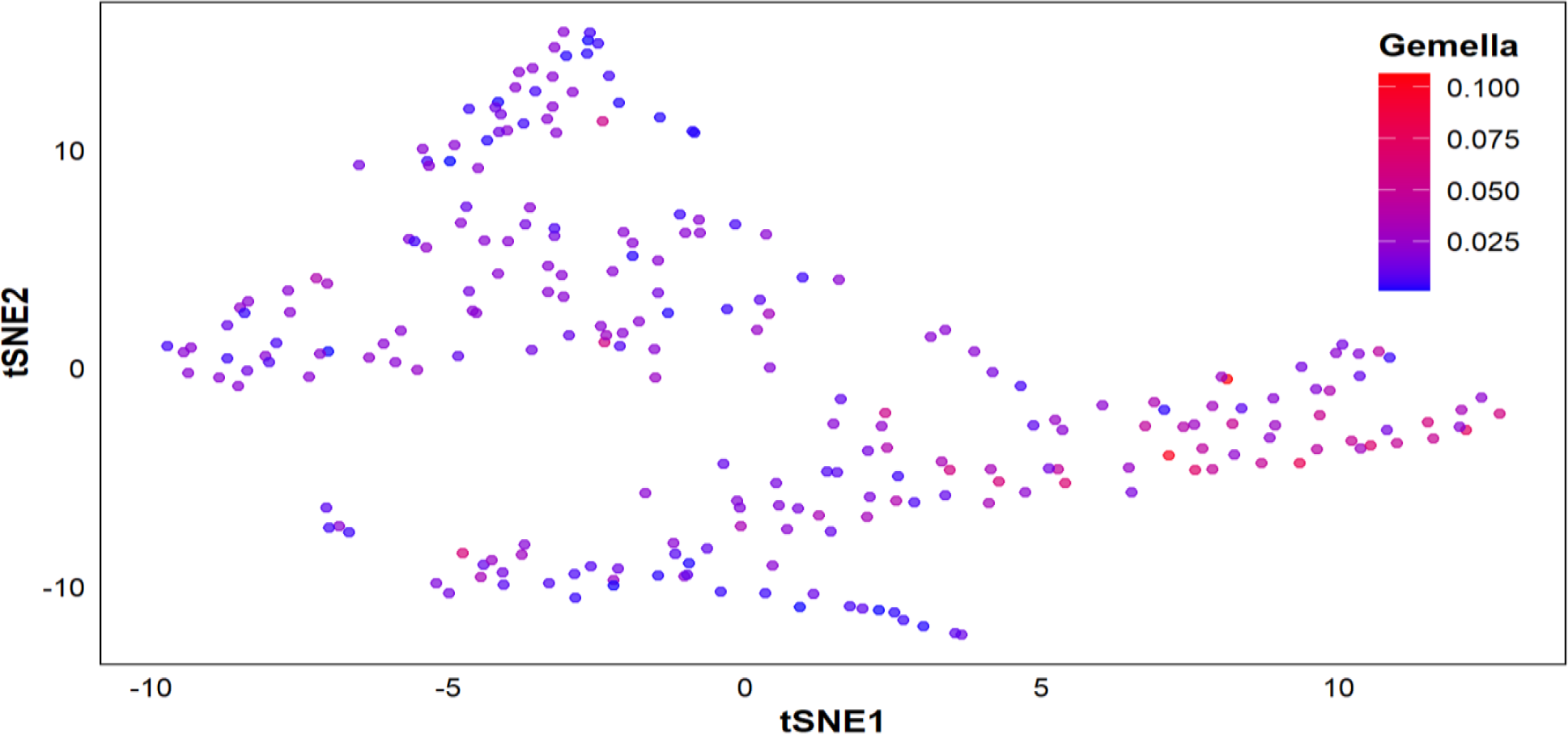
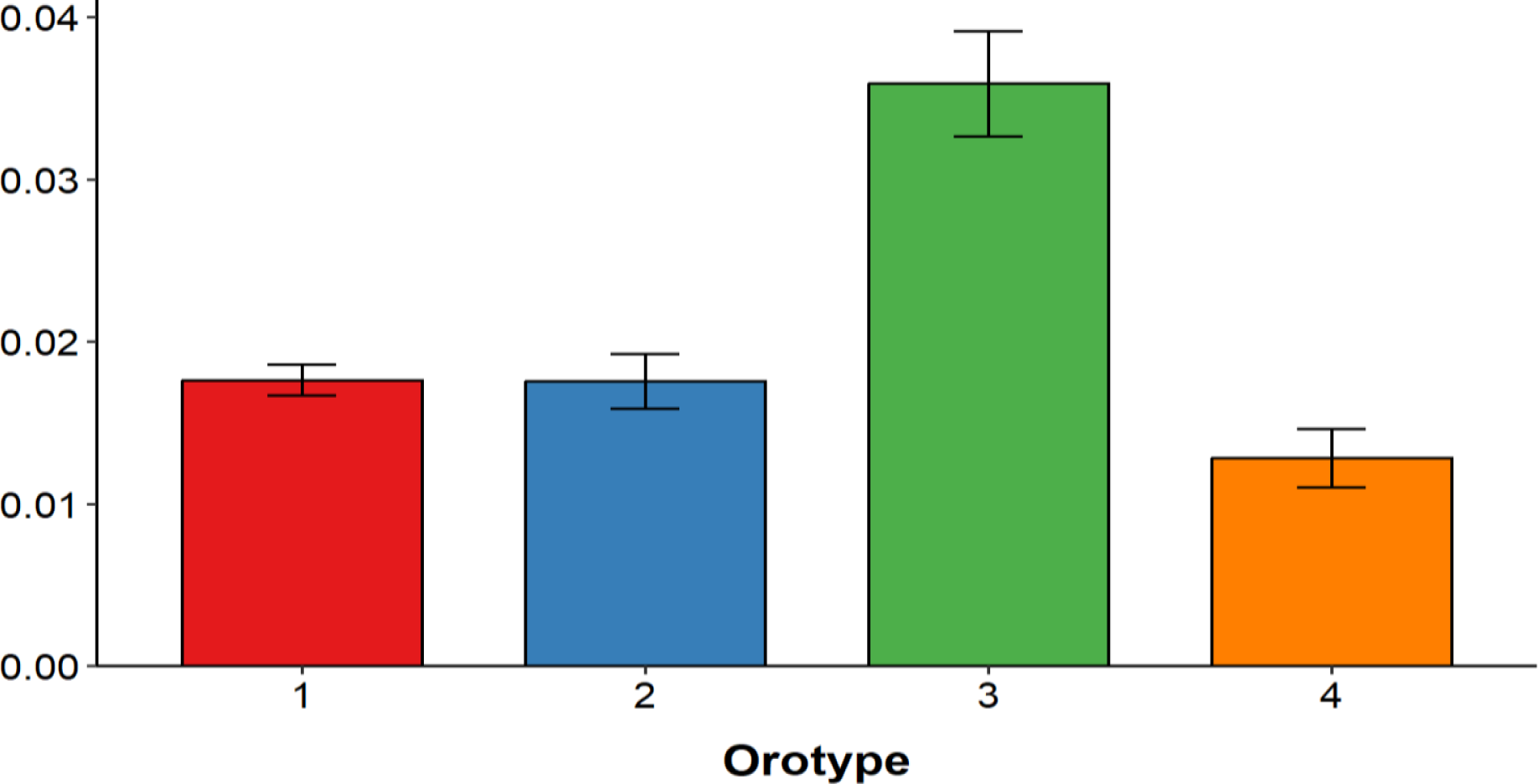
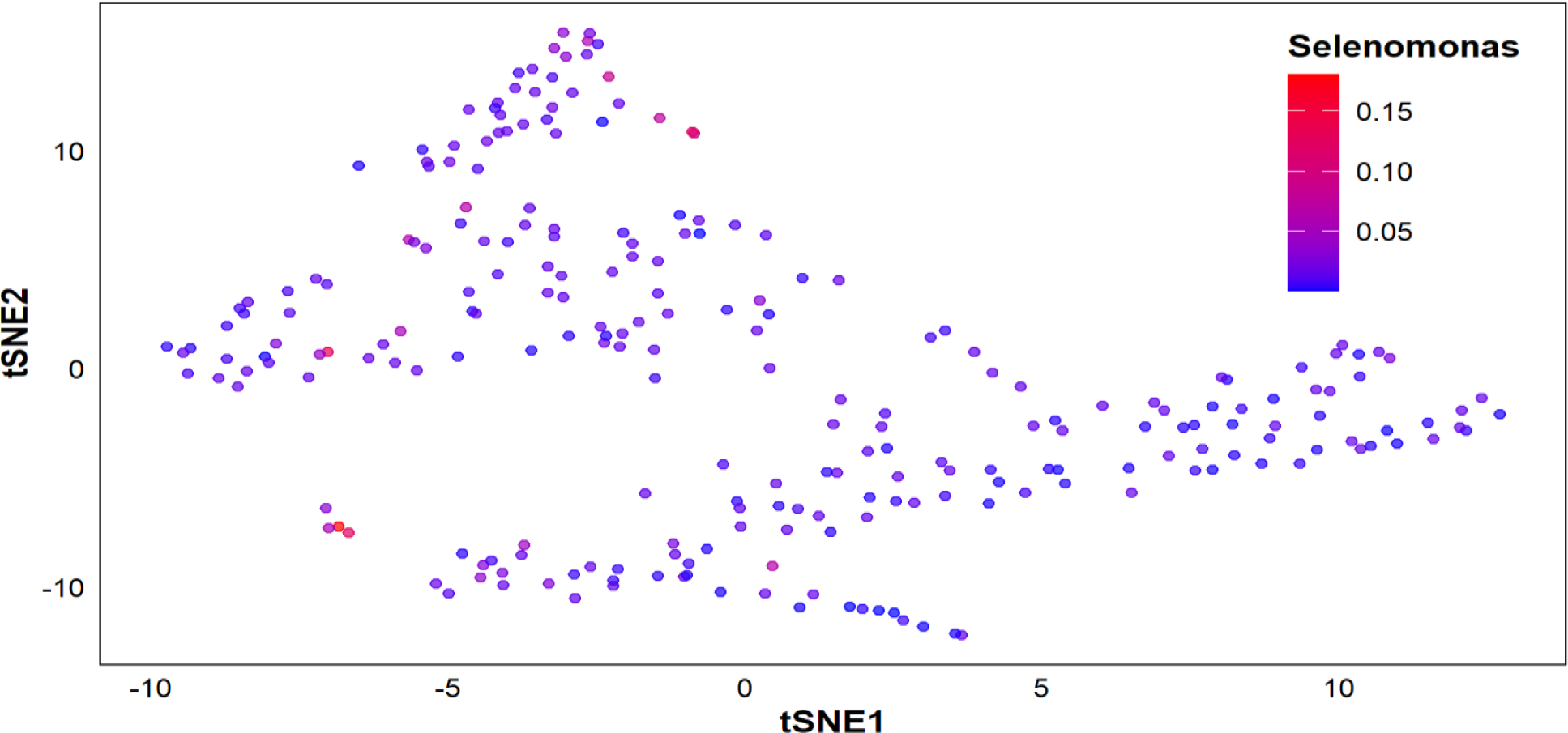
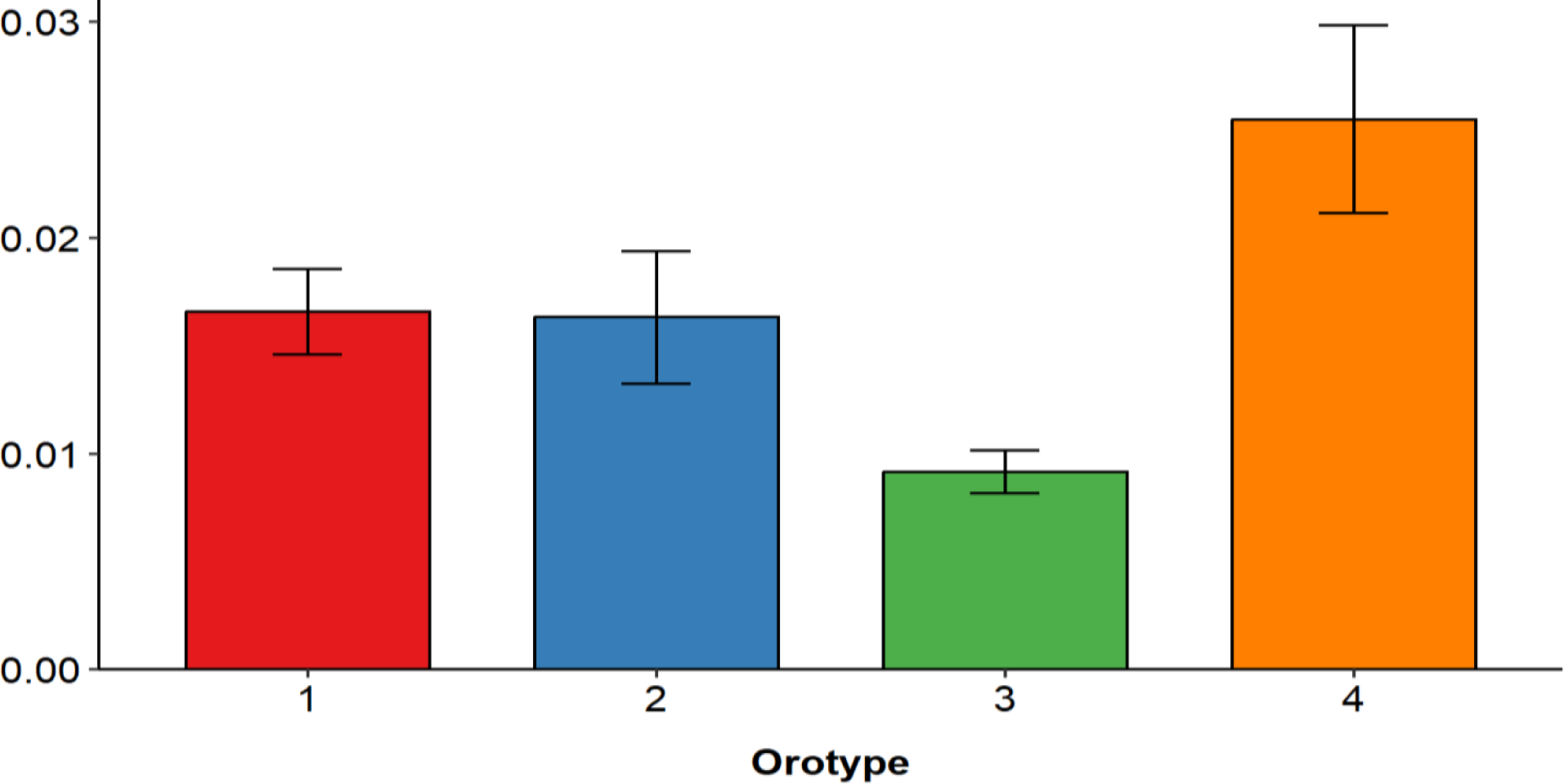
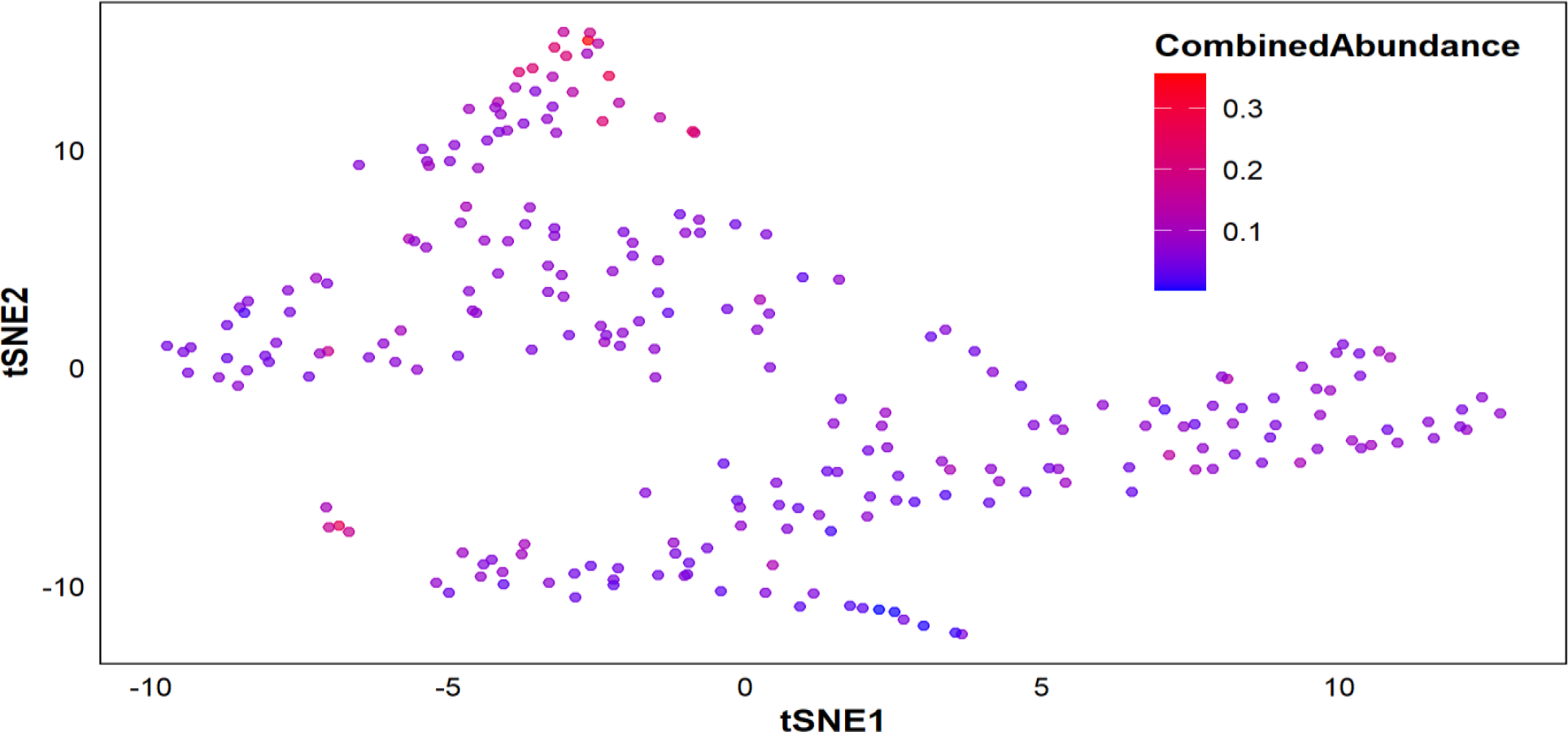
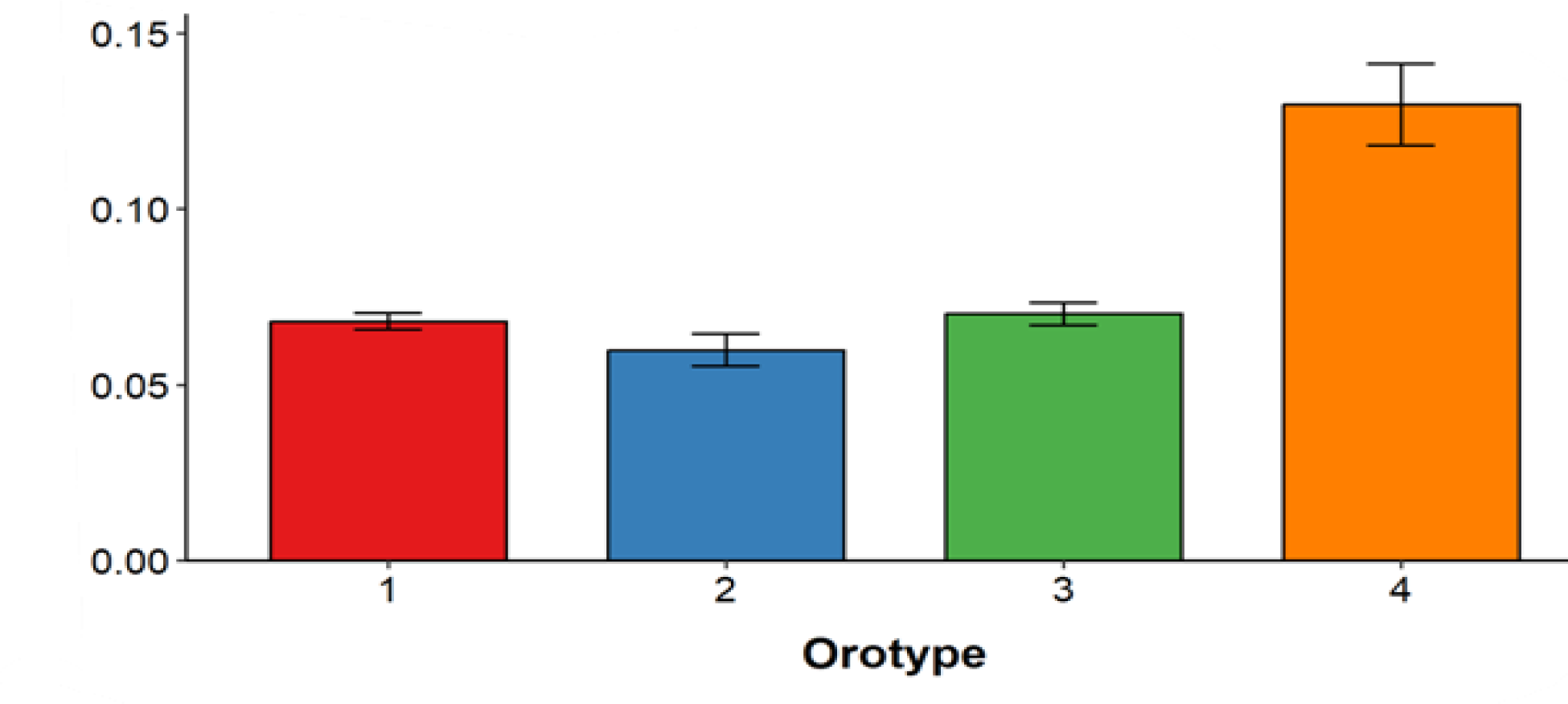
(a) The t-SNE plot is color-coded by the relative abundance of genus *Campylobacter*. (b) Mean and standard error of the relative abundance of genus *Campylobacter* in orotypes 1 to 4. *P* = 0.236 by Jonckheere-Terpstra trend test. (c)The t-SNE plot is color-coded by the relative abundance of genus *Gemella*. (d)Mean and standard error of the relative abundance of genus *Gemella* in orotypes 1 to 4. *P* = 0.652 by Jonckheere-Terpstra trend test. (e)The t-SNE plot is color-coded by the relative abundance of genus *Selenomonas*. (f)Mean and standard error of the relative abundance of genus *Selenomona* in orotypes 1 to 4. *P* = 0.859 by Jonckheere-Terpstra trend test. (g)The t-SNE plot is color-coded by the relative combine abundance of genus *Treponema, Gemella, Campylobacter, Selenomonas*. (h)Mean and standard error of the relative combine abundance of genus *Treponema, Gemella, Campylobacter, Selenomonas* in orotypes 1 to 4. p = 7.198e-05 by Jonckheere-Terpstra trend test.

**Fig S2:**
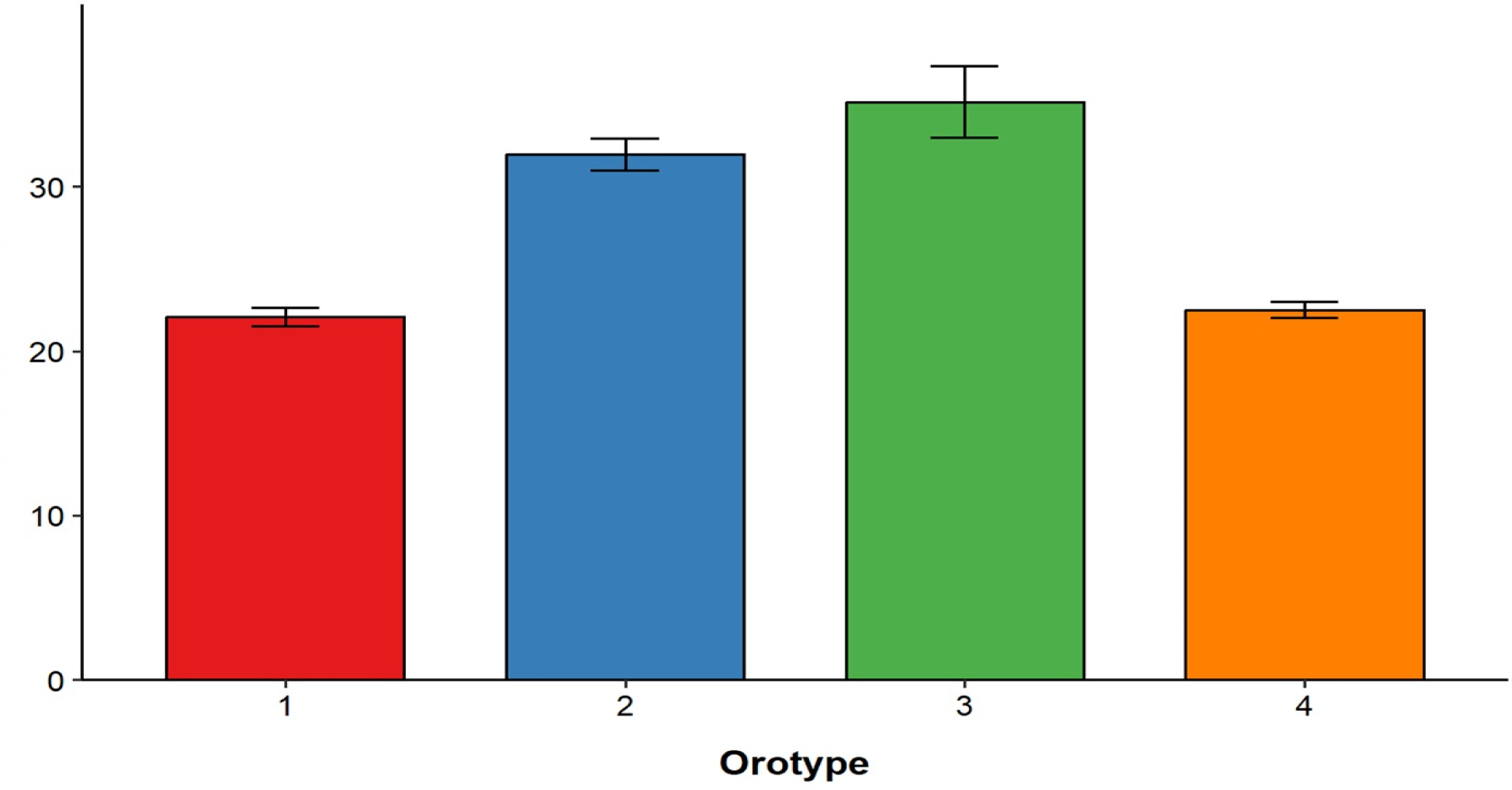
Mean and standard error of the Influenza mortality rates in orotypes 1 to 4. *p = 4e - 04* by Jonckheere-Terpstra trend test

**Supplementary Table 1:**
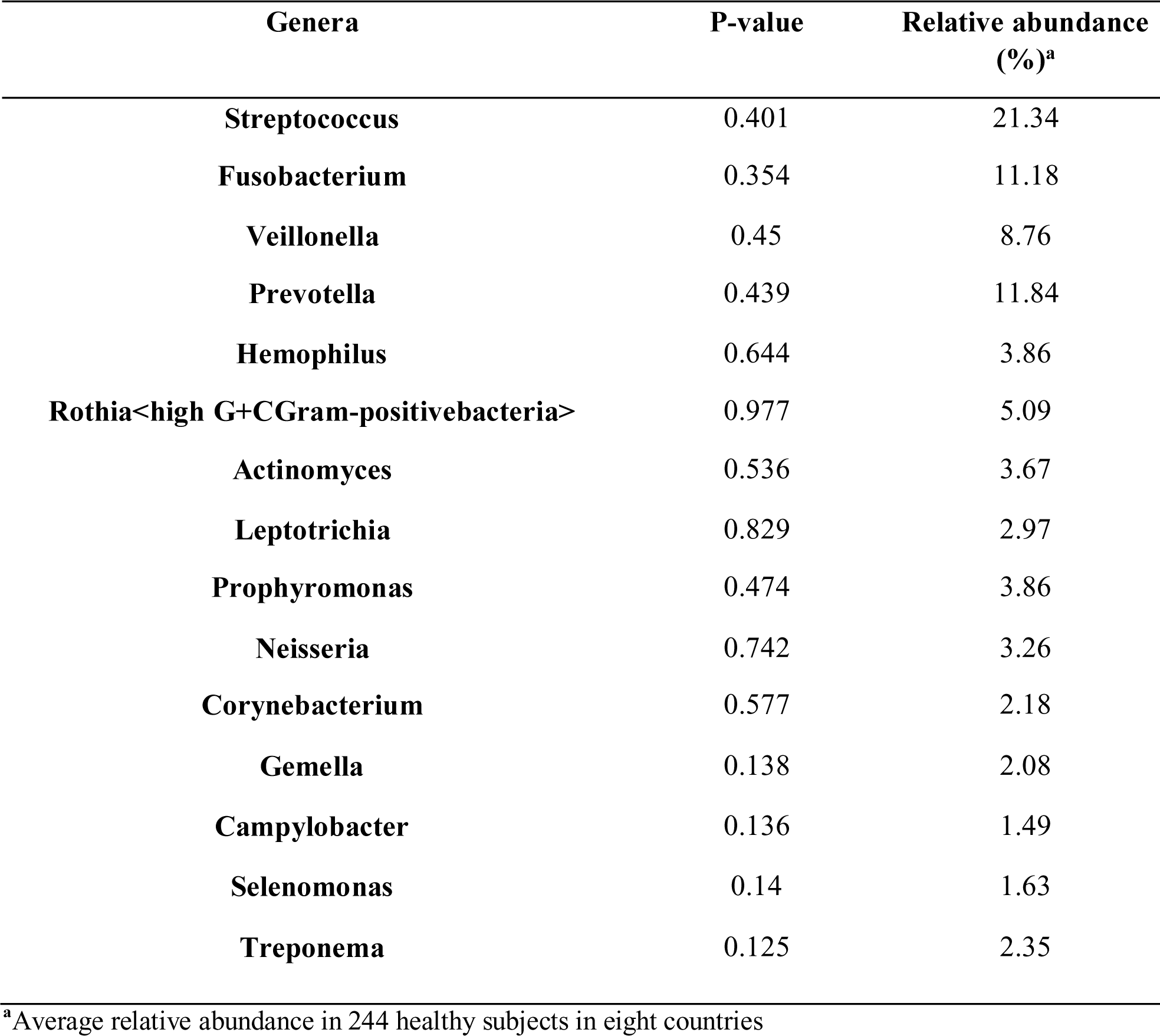
Generalized linear model (GLM) to predict the COVID-19 mortality rates with 15 oral bacteria.

**Supplementary Table 2:**
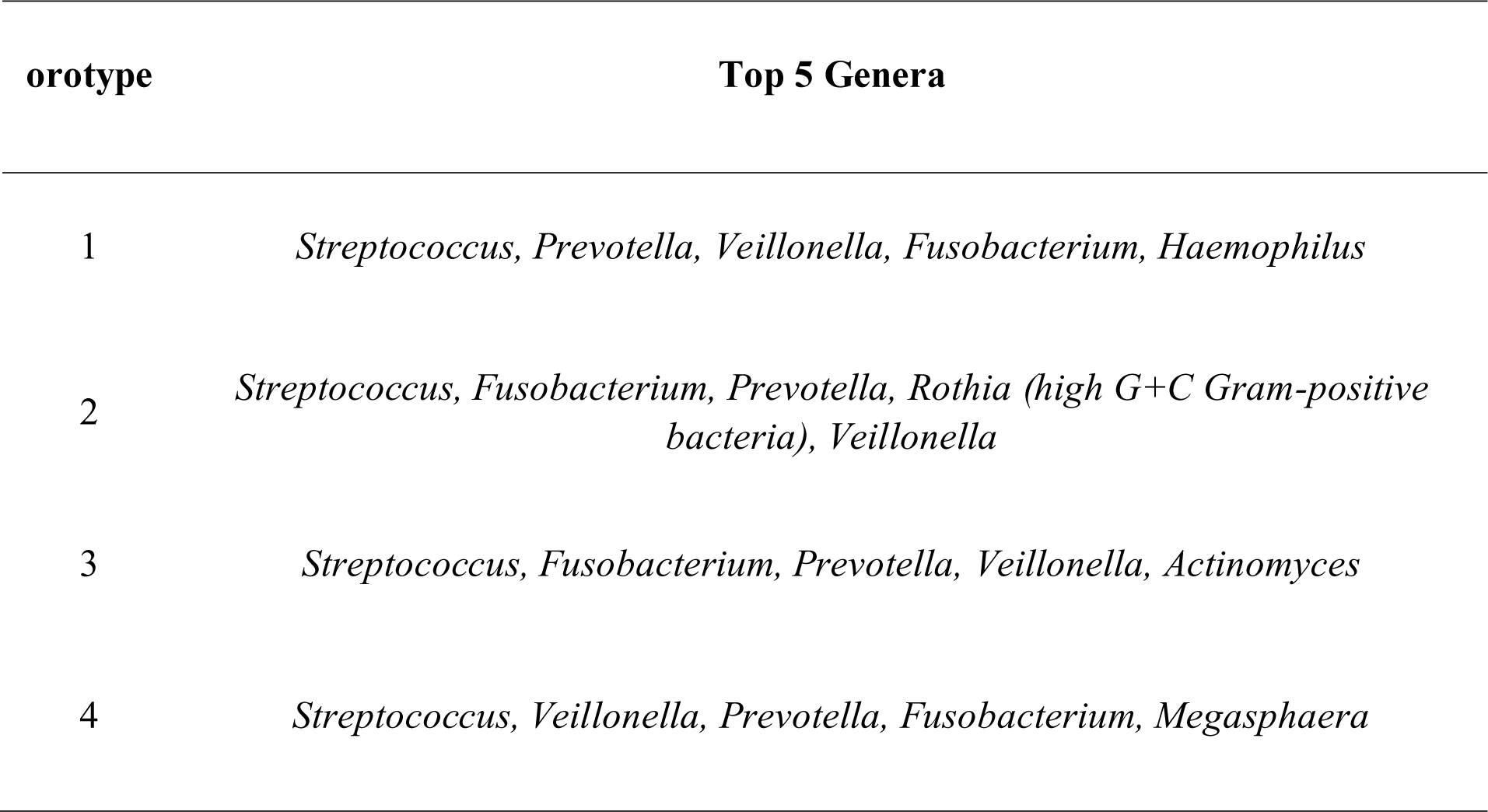
Most abundance genera in each orotype.

**Supplementary Table 3:**
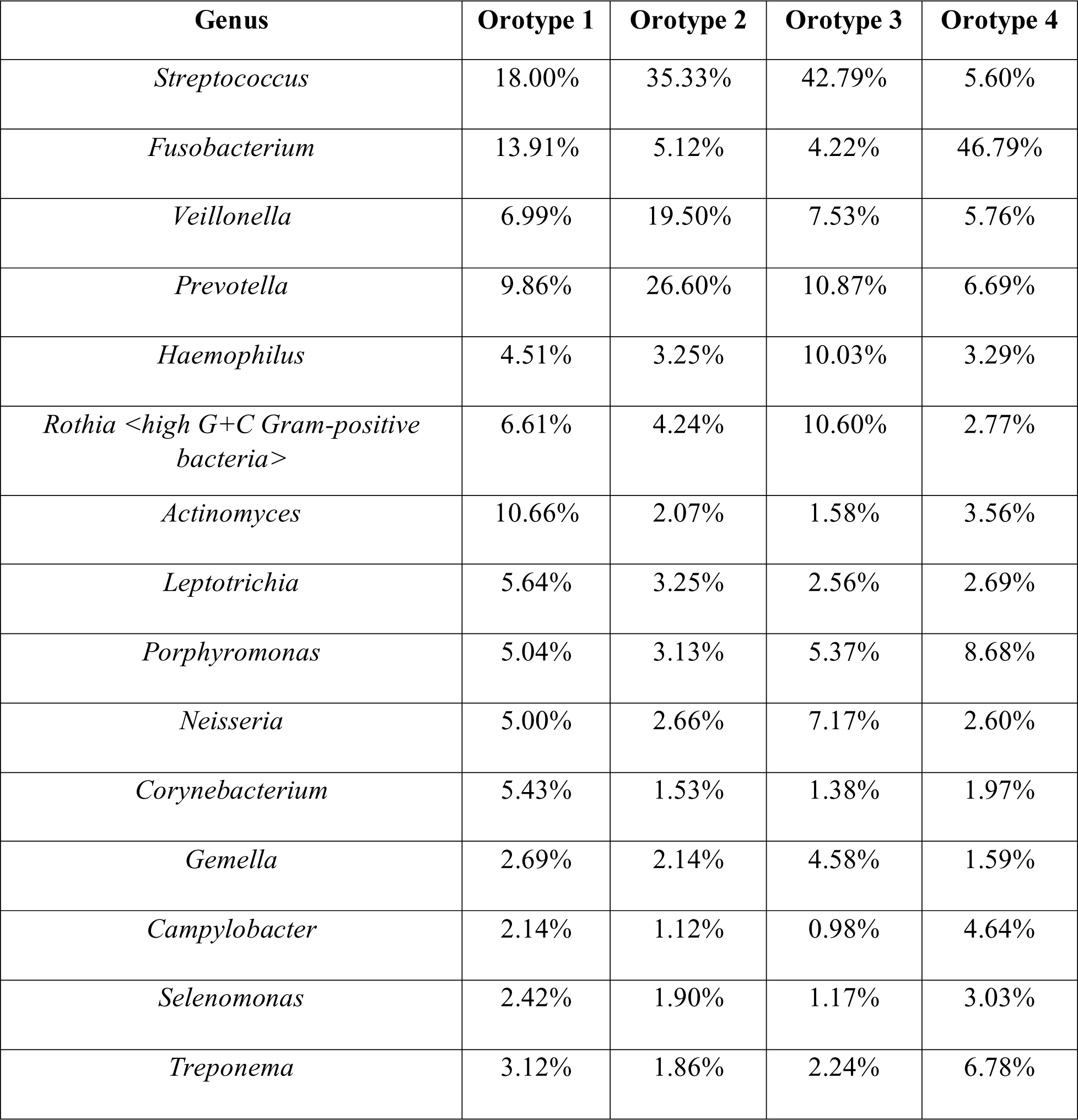
The mean relative abundances of 15 most prevalent genera for each orotype.

## Notes

### Competing Interest Statement

The authors have declared no competing interest.

### Funding Statement

This study did not receive any funding

### Author Declarations

we have taken this data from NCBI (https://www.ncbi.nlm.nih.gov/) and our world in data (https://ourworldindata.org/)

