## Supplementary figures and images for "Hydrogen Sulfide (H₂S)-Producing Oral Bacteria May Protect Against COVID-19"

### Fig S1a: The t-SNE plot is color-coded by the relative abundance of genus Campylobacter.

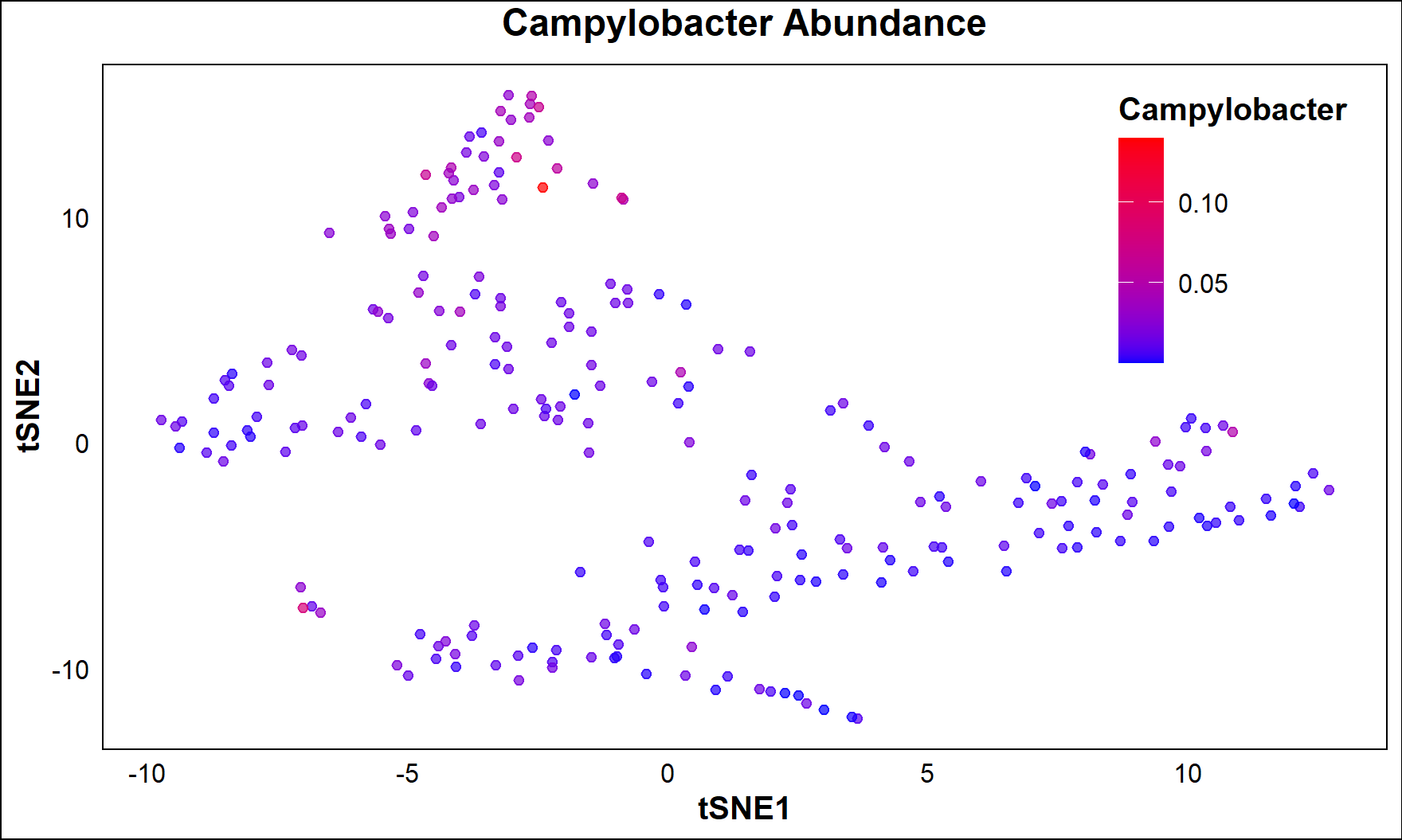

**Fig S1a: The t-SNE plot is color-coded by the relative abundance of genus *Campylobacter*.**

### Fig S1c: The t-SNE plot is color-coded by the relative abundance of genus Gemella.

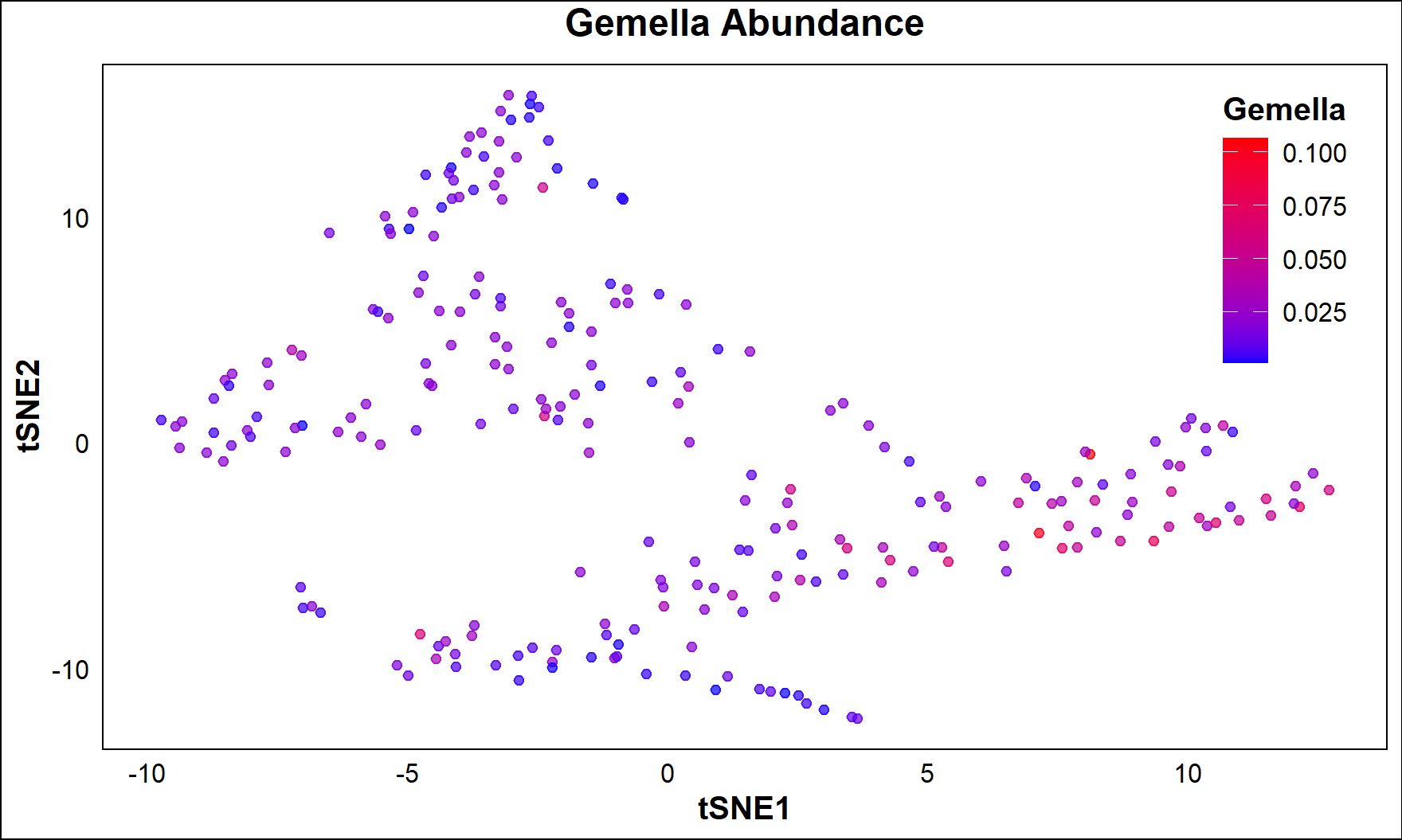

**Fig S1c: The t-SNE plot is color-coded by the relative abundance of genus *Gemella*.**

### Fig S1e: The t-SNE plot is color-coded by the relative abundance of genus Selenomonas.

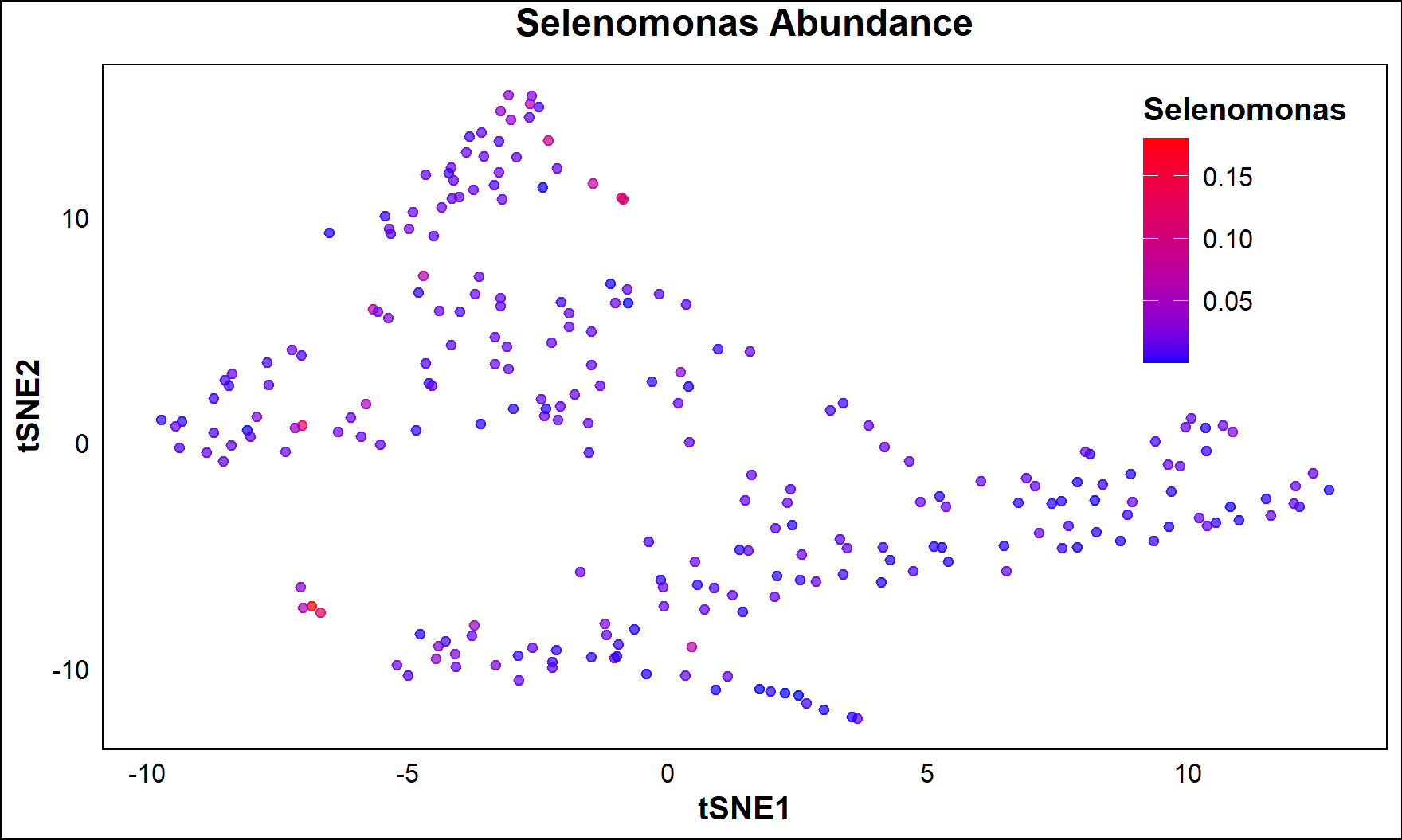

**Fig S1e: The t-SNE plot is color-coded by the relative abundance of genus *Selenomonas*.**
