## Supplementary material for "Hydrogen Sulfide (H₂S)-Producing Oral Bacteria May Protect Against COVID-19": Fig S1b: Mean and standard error of the relative abundance of genus Campylobacter in orotypes 1 to 4. P = 0.236 by Jonckheere-Terpstra trend test.

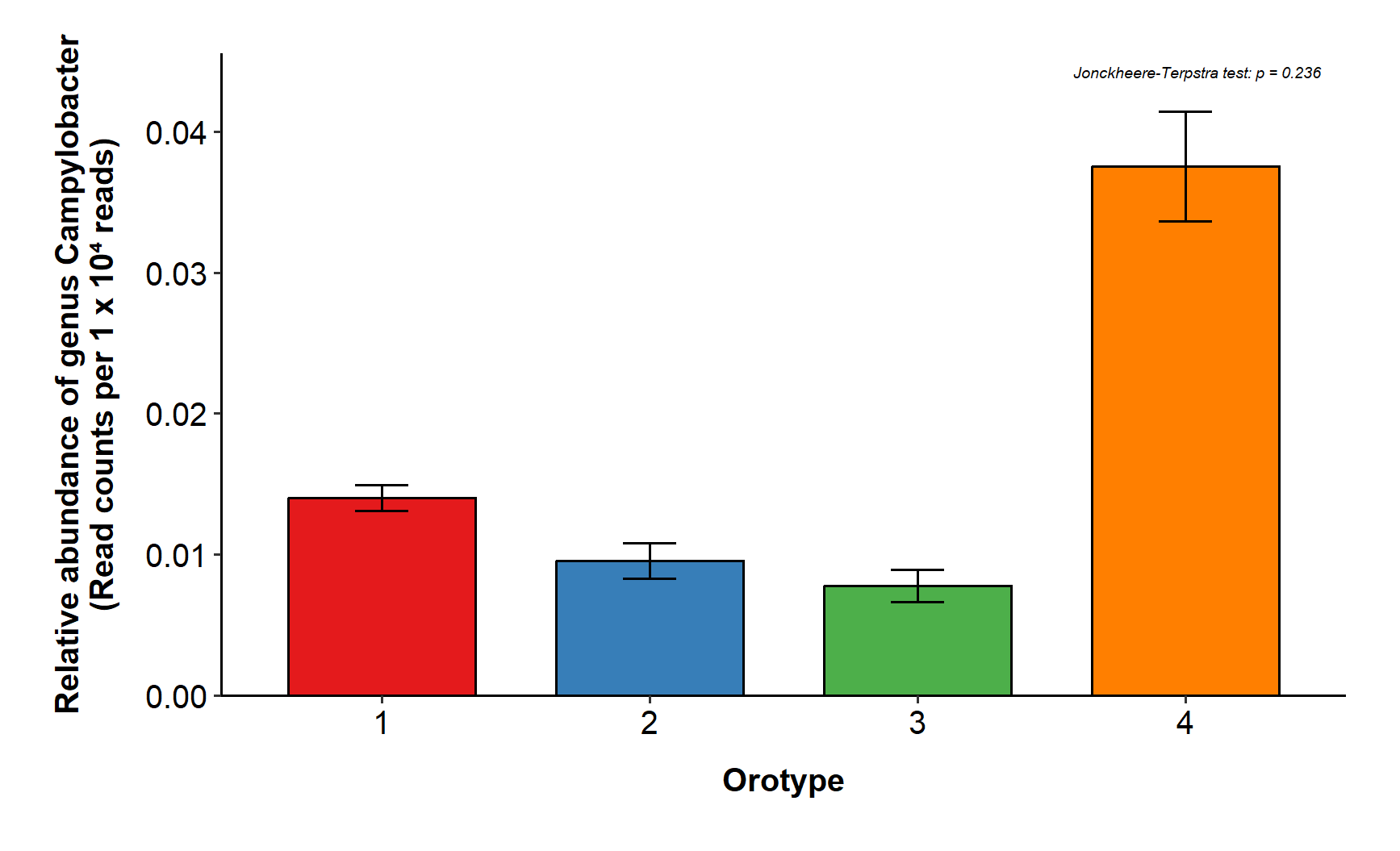

**Fig S1b:  Mean and standard error of the relative abundance of genus *Campylobacter* in orotypes 1 to 4. *P* = 0.236 by Jonckheere-Terpstra trend test.**
