## Supplementary material for "Hydrogen Sulfide (H₂S)-Producing Oral Bacteria May Protect Against COVID-19": Fig S1d: Mean and standard error of the relative abundance of genus Gemella in orotypes 1 to 4. P = 0.652 by Jonckheere-Terpstra trend test.

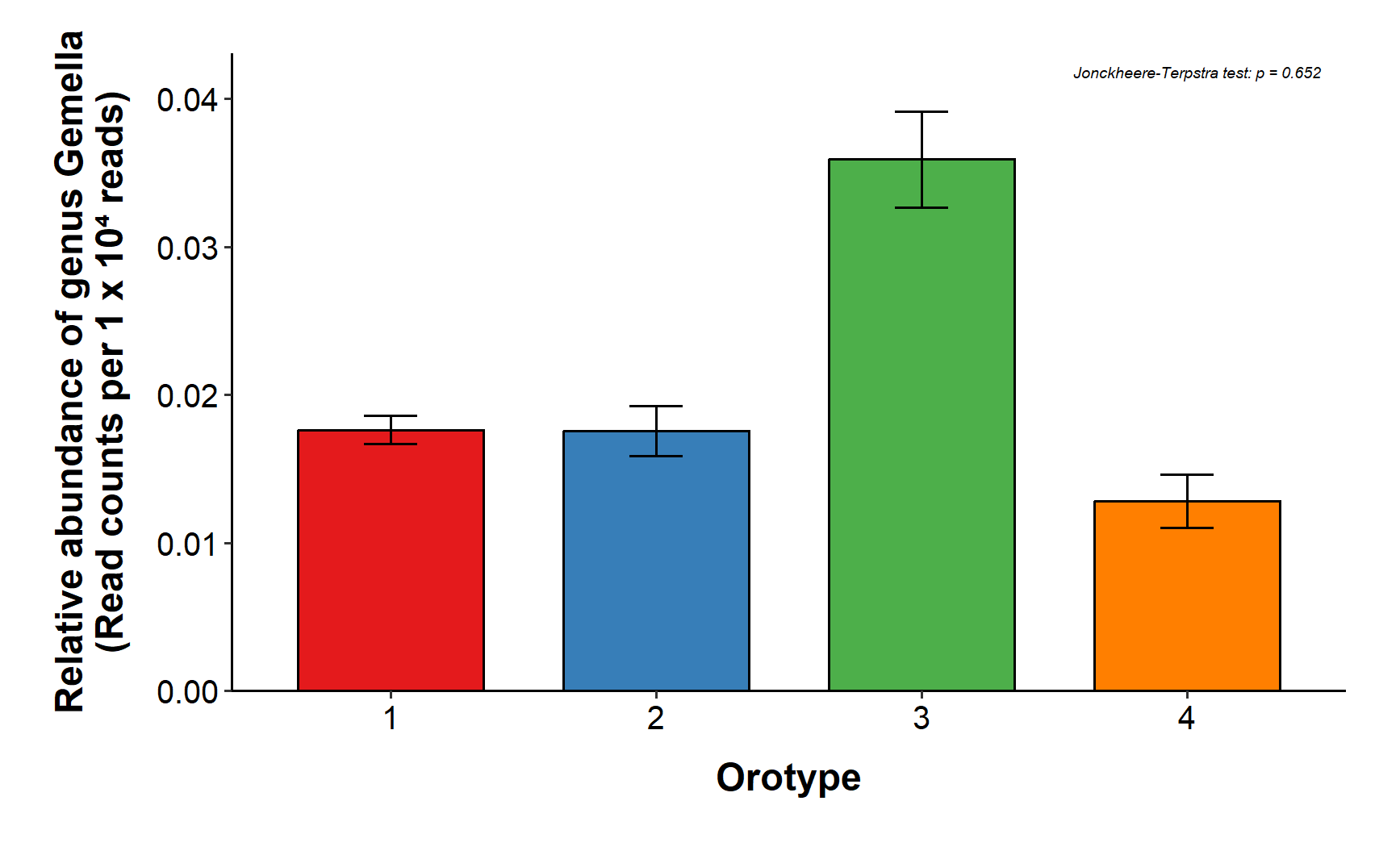

**Fig S1d: Mean and standard error of the relative abundance of genus *Gemella* in orotypes 1 to 4. *P* = 0.652 by Jonckheere-Terpstra trend test.**
