## Supplementary material for "Hydrogen Sulfide (H₂S)-Producing Oral Bacteria May Protect Against COVID-19": Fig S1g: The t-SNE plot is color-coded by the relative combine abundance of genus Treponema, Gemella, Campylobacter, Selenomonas.

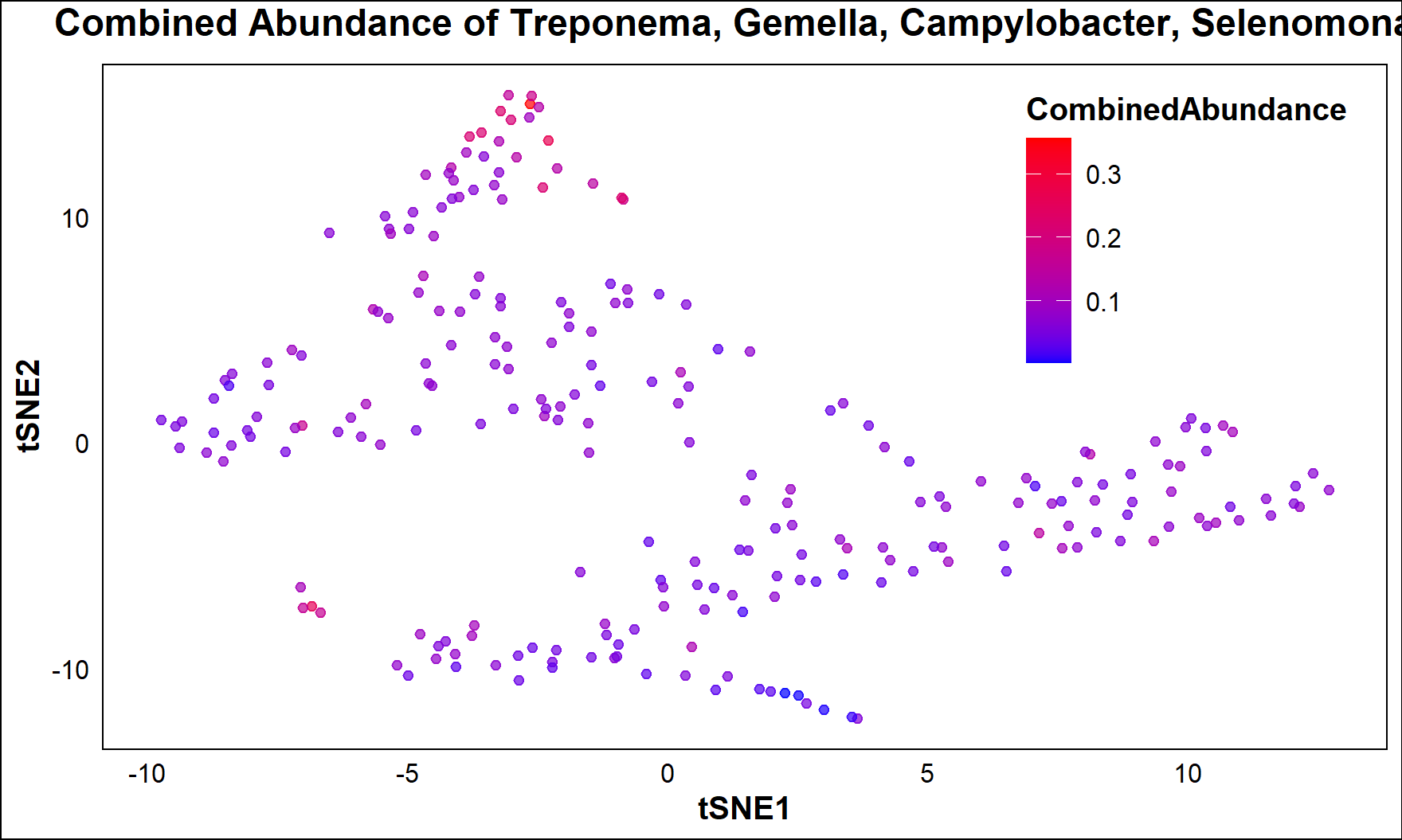

**Fig S1g: The t-SNE plot is color-coded by the relative combine abundance of genus *Treponema, Gemella, Campylobacter, Selenomonas*.**
