## Supplementary material for "Hydrogen Sulfide (H₂S)-Producing Oral Bacteria May Protect Against COVID-19": Fig S1h: Mean and standard error of the relative combine abundance of genus Treponema, Gemella, Campylobacter, Selenomonas in orotypes 1 to 4. p = 7.

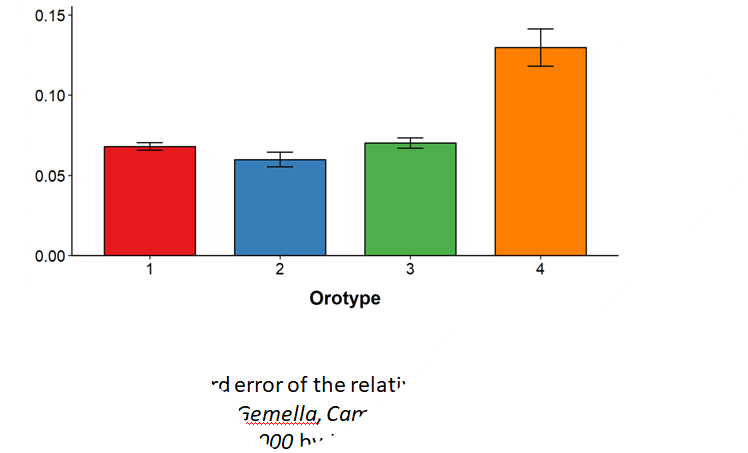

**Fig S1h: Mean and standard error of the relative combine abundance of genus *Treponema, Gemella, Campylobacter, Selenomonas* in orotypes 1 to 4.  p = 7.198e-05 by Jonckheere-Terpstra trend test.**
