## Supplementary material for "Hydrogen Sulfide (H₂S)-Producing Oral Bacteria May Protect Against COVID-19": Fig S2: Mean and standard error of the Influenza mortality rates in orotypes 1 to 4. p = 4e - 04 by Jonckheere-Terpstra trend test.

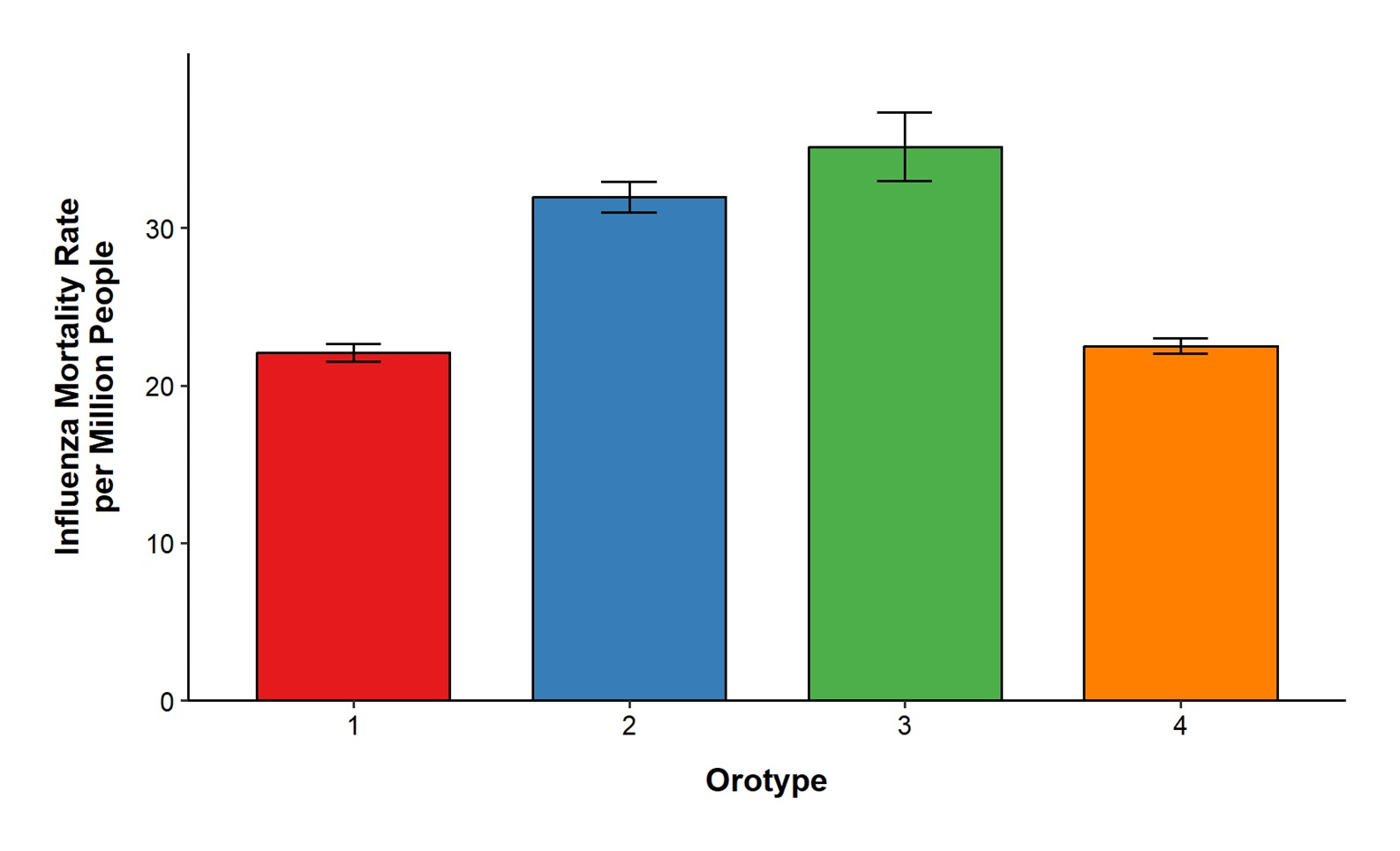

**Fig S2: Mean and standard error of the Influenza mortality rates in orotypes 1 to 4. *p = 4e - 04* by Jonckheere-Terpstra trend test.**
