## Supplementary Table 1: Generalized linear model (GLM) to predict the COVID-19 mortality rates with 15 oral bacteria for "Hydrogen Sulfide (H₂S)-Producing Oral Bacteria May Protect Against COVID-19"

| **Genera** | **P-value** | **Relative abundance (%)^a^** |
| --- | --- | --- |
| **Streptococcus** | 0.401 | 21.34 |
| **Fusobacterium** | 0.354 | 11.18 |
| **Veillonella** | 0.45 | 8.76 |
| **Prevotella** | 0.439 | 11.84 |
| **Hemophilus** | 0.644 | 3.86 |
| **Rothia<high G+CGram-positivebacteria>** | 0.977 | 5.09 |
| **Actinomyces** | 0.536 | 3.67 |
| **Leptotrichia** | 0.829 | 2.97 |
| **Prophyromonas** | 0.474 | 3.86 |
| **Neisseria** | 0.742 | 3.26 |
| **Corynebacterium** | 0.577 | 2.18 |
| **Gemella** | 0.138 | 2.08 |
| **Campylobacter** | 0.136 | 1.49 |
| **Selenomonas** | 0.14 | 1.63 |
| **Treponema** | 0.125 | 2.35 |

**^a^**Average relative abundance in 244 healthy subjects in eight countries
