## Supplementary Table 2: Most abundance genera in each orotype for "Hydrogen Sulfide (H₂S)-Producing Oral Bacteria May Protect Against COVID-19"

| **orotype** | **Top 5 Genera** |
| --- | --- |
| 1 | *Streptococcus, Prevotella, Veillonella, Fusobacterium, Haemophilus* |
| 2 | *Streptococcus, Fusobacterium, Prevotella, Rothia (high G+C Gram-positive bacteria), Veillonella* |
| 3 | *Streptococcus, Fusobacterium, Prevotella, Veillonella, Actinomyces* |
| 4 | *Streptococcus, Veillonella, Prevotella, Fusobacterium, Megasphaera* |
