## Supplementary Table 3: The mean relative abundances of 15 most prevalent genera for each orotype for "Hydrogen Sulfide (H₂S)-Producing Oral Bacteria May Protect Against COVID-19"

| **Genus** | **Orotype 1** | **Orotype 2** | **Orotype 3** | **Orotype 4** |
| --- | --- | --- | --- | --- |
| *Streptococcus* | 18.00% | 35.33% | 42.79% | 5.60% |
| *Fusobacterium* | 13.91% | 5.12% | 4.22% | 46.79% |
| *Veillonella* | 6.99% | 19.50% | 7.53% | 5.76% |
| *Prevotella* | 9.86% | 26.60% | 10.87% | 6.69% |
| *Haemophilus* | 4.51% | 3.25% | 10.03% | 3.29% |
| *Rothia <high G+C Gram-positive bacteria>* | 6.61% | 4.24% | 10.60% | 2.77% |
| *Actinomyces* | 10.66% | 2.07% | 1.58% | 3.56% |
| *Leptotrichia* | 5.64% | 3.25% | 2.56% | 2.69% |
| *Porphyromonas* | 5.04% | 3.13% | 5.37% | 8.68% |
| *Neisseria* | 5.00% | 2.66% | 7.17% | 2.60% |
| *Corynebacterium* | 5.43% | 1.53% | 1.38% | 1.97% |
| *Gemella* | 2.69% | 2.14% | 4.58% | 1.59% |
| *Campylobacter* | 2.14% | 1.12% | 0.98% | 4.64% |
| *Selenomonas* | 2.42% | 1.90% | 1.17% | 3.03% |
| *Treponema* | 3.12% | 1.86% | 2.24% | 6.78% |
